# A multi-ancestry genetic reference for the Quebec population

**DOI:** 10.1101/2025.05.14.25327536

**Authors:** Peyton McClelland, Georgette Femerling, Rose Laflamme, Alejandro Mejia-Garcia, Mohadese Sayahian Dehkordi, Hongyu Xiao, Alex Diaz-Papkovich, Justin Pelletier, Jean-Christophe Grenier, Ken Sin Lo, Luke Anderson-Trocmé, Justin Bellavance, Vincent Chapdelaine, Geneviève Gagnon, Annelie De Mori, Gerardo Martinez, Kristen Mohler, Thibault de Malliard, Catherine Labbé, Marjorie Labrecque, Alexandre Montpetit, Dan Spiegelman, Guy A Rouleau, Jean-François Théroux, Hufeng Zhou, Simon L Girard, Julie G Hussin, Anne-Marie Laberge, Claude Bhérer, Martine Tetreault, Sarah A Gagliano Taliun, Daniel Taliun, Simon Gravel, Guillaume Lettre

**Author notes:** Correspondence should be addressed to: Daniel Taliun, Simon Gravel and Guillaume Lettre. These authors contributed equally (Peyton McClelland, Georgette Femerling, Rose Laflamme, Alejandro Mejia-Garcia, Mohadese Sayahian Dehkordi). These authors co-supervised the work (Daniel Taliun, Simon Gravel, Guillaume Lettre).

## Abstract

While international efforts have characterized genetic variation in millions of individuals, the interplay of environmental, social, cultural, and genetic factors is poorly understood for most worldwide populations. The province of Quebec in Canada has been the site of numerous genetic studies, often focusing on individual Mendelian diseases in founder sub-populations. Here, we profiled and analyzed genome-wide genotyped variation in 29,337 Quebec residents from the large population-based cohort CARTaGENE (CaG), including rich phenotype and environmental data. We also sequenced the whole-genome of 2,173 CaG participants, including 163 and 132 individuals with grandparents born in Haiti and Morocco, respectively. We use this genetic information to gain insight into Quebec’s demography and to help interpret the potential significance of variants identified in clinically important genes. We built an imputation panel by phasing the CaG whole-genome sequence data and showed, using genome-wide association studies (GWAS), how it improves the discovery of phenotype-genotype associations in this population. We provide allele frequency information and GWAS results through dedicated and publicly available websites. The genetic data, paired with phenotypic and environmental information, is also available for research use upon scientific and ethical review.

## INTRODUCTION

The field of human genetics has benefited immensely from large national population-based cohorts such as deCODE, the UK Biobank, Biobank Japan, and FinnGen. While these cohorts offer unprecedented opportunities to improve statistical methods and discover new genotype-phenotype associations, they only capture a subset of the genetic and environmental variation that is susceptible to influence human populations in health and disease. Cohorts that are designed to represent populations with unique histories, environments, or geographic contexts can provide novel insights into the genetic architecture of human diseases, revealing population-specific variants, gene-environment interactions, and mechanisms of disease. They also can help tailor public health and precision medicine strategies (e.g. genetic screening programs or polygenic risk scores) using data that capture the unique characteristics of these populations.

The province of Quebec in Canada has a population of 8.8M individuals. Most Quebec residents derive ancestry from ∼8,500 individuals, primarily settlers from France, who moved to New France during the French colonial era in the 17^th^ and 18^th^ centuries^1^. Many genetic studies have studied Quebec residents with French-Canadian ancestry (QFC) to identify pathogenic variants that are relatively frequent in certain regions of Quebec but very rare elsewhere^2–4^. Collectively, Quebec residents have ancestry from the Americas (especially among Indigenous individuals), Europe (e.g., early French and British colonists, or more recent immigrants), as well as regions around the world primarily via more recent immigration waves (e.g. from South Asia, Northern Africa or the Caribbean). Genetic ancestry is not distributed uniformly across the population but correlates with geographic, ethnic and linguistic characteristics of individuals, among other factors^2,5–7^. To provide a comparative framework for analyzing genetic patterns, we use population descriptors throughout this study that aim to capture geographical differences among ancestors of participants (see glossary in **Supplementary Notes**), although we acknowledge that these labels do not account for the complexity of human ancestral relationships.

CARTaGENE (CaG) is the largest population-based cohort in Quebec. It was initiated in 2007 to facilitate genetic and epidemiological research in the cosmopolitan population of the province^8^. This cohort includes 43,032 participants (55% women) that were recruited from six different metropolitan areas of Quebec, including Montreal (70% of the cohort), Quebec City (15.3%), Sherbrooke (4.5%), Saguenay (4.2%), Gatineau (3.7%) and Trois-Rivières (2.3%). Approximately 16% of participants self-identified as having non-European ethnicity. Participants were 40-69 years old at the baseline visit and provided extensive health and socio-demographic information. They are being followed regularly through recontact phases and ancillary studies, and it is possible to link each participant with their corresponding health record through the provincial healthcare system and their environmental data through the Canadian Urban Environmental Health Research Consortium (CANUE). The genealogies of 7,894 CaG participants were reconstructed by the BALSAC Project, a vast reconstruction of Quebec genealogy that is especially complete among QFC individuals^9^. CaG also has a biobank with biospecimens (e.g. DNA, blood, plasma) available for 29,337 participants.

Here we present analyses of genome-wide genotyping array data for 29,337 CaG participants, showing how historical and demographic events such as waves of immigration have shaped the genetic structure of modern Quebec. We generated whole-genome sequence (WGS) data from 2,173 CaG participants, and showed how this resource facilitates the study of rare genetic diseases as well as complex traits through improved genotype imputation of single nucleotide variants (SNVs), insertions-deletions (indels), structural variants (SVs) and human leukocyte antigen (HLA) alleles. We conducted genome-wide association studies (GWAS) for 42 quantitative phenotypes, identifying >350 independent loci (in participants of European ancestry, >450 loci in cross-ancestry GWAS) with variants that reach genome-wide significance (P-value <5×10^−8^). We released CaG allele frequency information, and GWAS and expression quantitative trait loci (eQTL) results through publicly available CaG browsers. Individual-level data from the CaG dataset is available to both academic and industry-sponsored research projects following scientific and ethical review.

## RESULTS

### Genetic structure in CaG

Genome-wide genotyping array data on the Illumina GSA platform is available for 29,337 participants who consented for DNA research (**Supplementary Table 1**). Principal component (PC) analysis captures main axes of genetic variation in CaG. The first and second PCs capture continental genetic ancestry patterns (**Fig. 1A**), and the third and fourth PCs distinguish a branch corresponding to a well-known founder event in the regions of Charlevoix and Saguenay-Lac-St-Jean (SLSJ) in Quebec, a cline with individuals of Mediterranean ancestries (North Africa and Middle Eastern), and a branch with ancestry from Latin America (**Supplementary Fig. 1A**). Uniform manifold approximation and projection (UMAP)^10^ of the first ten PCs is characterized by clusters often correlating with country of origin (**Fig. 1B** and **Supplementary Fig. 1**). Results from identity-by-descent (IBD) segment analyses (**Methods**) were in line with the PC-based genetic similarity patterns, but helped reveal finer clusters of participants (**Supplementary Notes**, **Supplementary Fig. 2-3**). Among QFC, clusters primarily reflect geographical variation, including the previously described founder effect from the SLSJ region (**Supplementary Fig. 1**)^5,11^. Both IBD and UMAP also revealed a group of participants with ancestors from the Gaspé Peninsula or the Canadian Atlantic provinces, presumably related to the Acadian population^11,12^.

**Figure 1.**
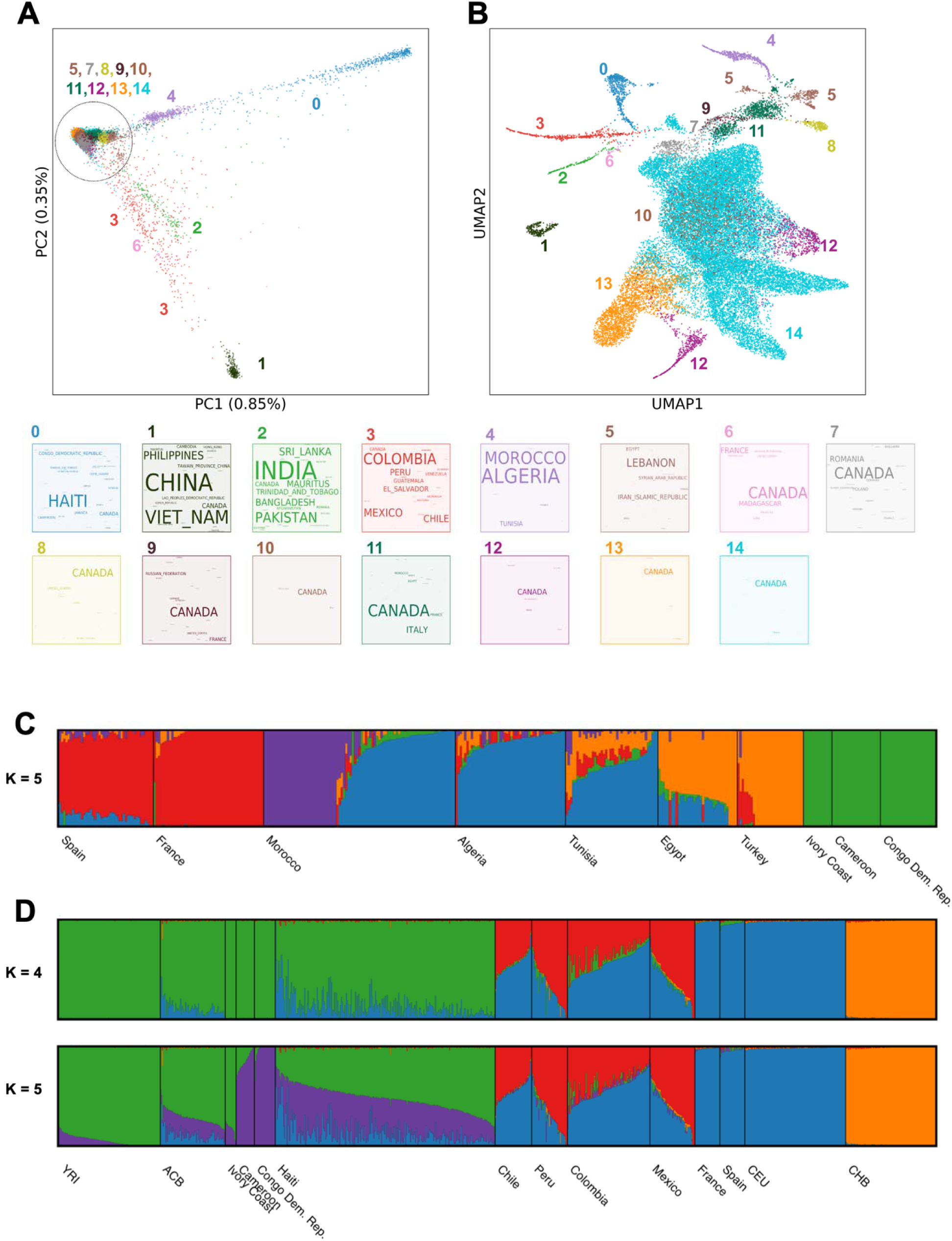
Genetic structure in the CaG population-based cohort of Quebec. (**A**) Principal components 1 and 2, labelled and colored by HDBSCAN clusters, as described in **Methods**. Word clouds of the countries of birth are provided for each cluster. Global population structure within the cohort is shaped by a combination of domestic structure and recent migration. (**B**) UMAP of the top 10 principal components of CaG, labelled and colored by the same HDBSCAN clusters as in **A**. Clusters reflect recent migration history, admixture, and domestic genetic structure within Quebec. Some clusters, such as 10 and 12, are not evident in the 2D UMAP but form in the higher dimensional UMAP used in HDBSCAN. Word clouds for other variables are available in **Supplementary Fig. 1**. (**C-D**) Estimation of admixture proportions among CaG participants whose four grandparents were born in Morocco (QMO, **C**) or Haiti (QHA, **D**). ADMIXTURE estimation was run independently for each group for varied number K of components (**Methods**). Reported Ks were chosen by cross-validation analysis (**Supplementary Fig. 4-5** for all Ks). The plots show CaG individuals with four grandparents born in same country, and three additional populations from the 1000 Genomes Project as reference in (**D**): YRI: Yoruba in Nigeria, CHB: Han Chinese in Beijing, China, ACB: African Caribbean in Barbados.

### Migrations that shaped Quebec

Ancestral component analysis in CaG reflected a range of demographic effects related to human migrations. Admixture analysis of participants born in North Africa showed a longitudinal gradient of ancestry across North Africa, consistent with back-to-Africa migrations (**Fig. 1C** and **Supplementary Fig. 4A**)^13^. The analysis also reflected the more recent waves of migration to Quebec. For example, the 201 individuals with four Morocco-born grandparents (QMO) form two clusters (**Fig. 1C**), which correlate with self-reported ethnicity variables. Cluster 1 (primarily blue) has “Arab” as modal self-reported ethnicity, whereas cluster 2 (primarily purple) has “Jewish” as modal self-reported ethnicity. Most QMO participants in CaG were themselves born in Morocco (97%) and reported either the year of immigration to Quebec or the age at which they immigrated. QMO individuals in cluster 2 are born earlier on average (mean=1951, standard deviation [SD]=7.6) compared to individuals in cluster 1 (mean=1962, SD=6.5). The average year of immigration differed between clusters (cluster 1, mean=1998 and SD=9; cluster 2, mean=1977, SD=11) and the reported ethnicity in each cluster reflects the two historically documented waves of Moroccan immigration to Quebec (**Supplementary Fig. 4B**)^14,15^. In contrast to older individuals, QMO individuals born after 1965 in cluster 2 are more likely to be children of migrants, as the majority were born outside of Morocco (5 out of 7). Therefore, any focused analysis of the QMO will require careful consideration of confounding between age, genetic ancestry, generation status, and ethnicity.

Haiti is another important source of immigration in Quebec. The distribution of immigration year (mode=1974, mean=1986, SD=13) among 267 individuals with four Haiti-born grandparents (QHA) reflects the increase of immigration from Haiti to Quebec in the early 1970s, referred to as the “second wave”, which formed the largest immigration group in Quebec between 1973 and 1974 (**Supplementary Fig. 5**)^16,17^. At the continental level, we find appreciable variation in estimated European and African ancestry proportions, and limited ancestry from the Americas (from K=4, mean AMR=0.5%, EUR=13.3% and AFR=86.2%, SD for AFR=13.2%) (**Fig. 1D**). The proportion of European ancestry in QHA individuals is comparable if marginally lower than the value of 16% reported previously in a small sample of Haiti-born individuals in South Florida^18^. At K=5, the African ancestry component separates in two, with a purple component predominant in CaG individuals born in Cameroon and Democratic Republic of the Congo, and a green component predominant in Yoruba individuals from Ibadan, Nigeria (YRI) and CaG individuals from Ivory Coast. In this model, QHA individuals are represented as having comparable levels of both ancestries, in contrast with Afro-Caribbean individuals from Barbados (ACB) from the 1000 Genomes project, which have more Yoruba-like (green) ancestry.

### Ancestry from the Americas

The original inhabitants of Quebec were the First Nations and Inuit peoples. Ancestry from the Americas prior to European settlement is present at different levels in the general population and is relevant for the prevalence of variants present in the First Peoples of Quebec^3,19,20^. The CaG cohort was recruited from individuals enrolled in the provincial healthcare plan. While this plan covers most individuals in the province, healthcare in Indigenous populations follows a few different models and therefore Indigenous individuals enrolled in CaG are not statistically representative of the overall Indigenous population of the province of Quebec. Out of 500 randomly selected CaG participants with four grandparents born in Canada, the proportion of estimated ancestry from the Americas was 0.4%, from K=4, with no individual carrying more than 5% indigenous ancestry (**Supplementary Fig. 6**).

### Variant discovery in CaG using high-depth whole-genome DNA sequencing

To complement the genotyping array data and identify rare variants found in the Quebec population, we performed high-coverage WGS of 2,184 CaG participants, including 1,765 QFC, 163 QHA participants, and 132 QMO participants (the remaining 124 participants were non-QFC European-ancestry individuals) (**Supplementary Fig. 7** and **Supplementary Table 1**). After quality-control, we obtained WGS data for 2,173 participants at a mean coverage of 33.4X (1,756 QFC, 163 QHA, 132 QMO). In total, we identified 80,407,530 variants (16.8% novel based on dbSNP v154), including >3.1M singletons (**Supplementary Table 2**). While the QHA and QMO samples accounted for 14% of our WGS effort, they yielded 22% of the newly discovered variants (**Supplementary Fig. 8**). To enable fast variant allele frequency queries and support clinical genetic applications, we have released allele frequencies for each of the three main groups (QFC, QHA, QMO) sequenced in CaG through a publicly available CaG variant browser (https://cartagene-bravo.cerc-genomic-medicine.ca/). We expect that the data in this browser will facilitate the interpretation of variants discovered in patients of French-Canadian, Haitian, and Moroccan genetic ancestries in Quebec and elsewhere.

From the WGS data, we identified high-impact coding variants, defined as potentially functional (annotated as nonsense, frameshift or essential splice site) and/or pathogenic (based on ClinVar), and compared their frequency with the frequency in the best matched population from gnomAD v4 (**Supplementary Fig. 9** and **Supplementary Table 3**)^21^. As expected, given the founder events in the QFC population history, the allele frequencies of many clinically important rare variants are higher in CaG participants than in gnomAD non-Finnish Europeans (NFE) (**Supplementary Table 4**). Although the sample size is limited, we also recovered ClinVar-annotated pathogenic variants in the QHA and QMO WGS subsets that are more frequent than in the matched gnomAD populations, such as a variant in *AGPAT2* (causing lipodystrophy) previously found in families of African descent, and variants in *FAM161A* (causing retinitis pigmentosa) and *DSG4* (causing hypotrichosis) previously identified in Moroccan Jewish families (**Supplementary Table 4**).

### Structural variants

From the WGS data, we identified and genotyped 25,820 SVs, including 24,022 deletions and 1,798 duplications (**Supplementary Fig. 10A**). On average, an individual CaG genome includes 4,118 SVs (**Supplementary Fig. 10B**). When we compared with the gnomAD-SV database (allele frequencies from all gnomAD populations)^22^, we found that 74% of the CaG SVs were in the database with highly concordant allele frequencies (Pearson’s *r*=0.81 for deletions and *r*=0.54 for duplications, **Supplementary Fig. 10C**). Most SVs are rare (80% have a minor allele frequency [MAF] <5%) and encompass on average 10.5-kb of genomic DNA sequence (median length 1.3-kb, **Supplementary Fig. 10D**-E). The majority (∼70%) of the SVs were specific to an ancestral group, including 14.0% and 4.2% that were only found in QHA and QMO participants, respectively (**Supplementary Fig. 10F-G).** Using AnnotSV version 3.2.3, we annotated the CaG SVs: 46.5% overlap a gene, and 107 are predicted to be pathogenic (class 5, AnnotSV ranking score ≥0.99, based on the joint consensus recommendation of American College of Medical Genetics (ACMG) and ClinGen) (**Supplementary Fig. 10H** and **Supplementary Table 5**). For instance, we found one carrier of a 7.9-kb deletion encompassing the promoter and first exon of the *HEXA* gene: this deletion has been reported in QFC patients with Tay-Sachs disease^23^. We also identified six carriers of the 15+kb *LDLR* deletion, which is known to cause 60% of familial hypercholesterolemia cases in QFC^24^. In the CaG WGS dataset, the 15+kb *LDLR* deletion is associated with an increase in plasma LDL-cholesterol levels (+1.45 mmol/deletion, P-value=4.8×10^−5^).

### HLA genotyping

From the WGS data, we performed HLA allele genotyping for 11 classical class I and class II HLA genes *(HLA-A, -B, -C, -DPA1, -DPB1, -DQA1, -DQB1, -DRB1, -E, -F,* and *-G)* to G-group resolution using the software HLA*LA^25^. After variant- and sample-level quality filtering, we genotyped 43,189 HLA alleles, on average 0.9 per gene and per haploid genome. We observed 307 distinct HLA alleles, distributed as follows: 55 *HLA-A*, 73 *-B*, 38 *-C*, 9 *-DPA1*, 26 *-DPB1*, 11 *-DQA1*, 17 *-DQB1*, 55 *-DRB1*, 5 *-E*, 2 *-F*, and 16 –*G* (**Supplementary Fig. 11A**). For 57 one-field HLA alleles present in CaG and a reference dataset for Quebec^26^, allele frequencies correlated well (Pearson’s *r* = 0.97) (**Supplementary Fig. 11B** and **Supplementary Table 6**). Regional allele frequencies across Quebec were also well correlated, with Pearson’s *r* ranging from 0.86-0.96, demonstrating representativeness and confidence of HLA allele calling in CaG (**Supplementary Table 6**). Cohort-wide and population-specific HLA allele frequencies were compared to the largest publicly available multi-ancestry HLA reference panel (N=21,546)^27^. 18 HLA alleles from the CaG dataset were not present in this global reference panel, and an additional 89 alleles were at significantly different frequencies (Fisher’s exact test P-value <2.3×10^−4^). Frequencies remained significantly different for 51 alleles when tested in the QFC CaG subset as compared to European frequencies in the reference panel (**Supplementary Fig. 11C** and **Supplementary Table 7**). Our analysis identified one allele of possible interest, the HLA-A*24:02:02 allele (allele frequency [AF]=0.0023 in CaG, AF=2.3×10^−5^ in global reference panel, P-value=4.2×10^−8^), which is associated with increased risk of adverse drug reactions^28,29^ and was one of the most significantly enriched alleles in CaG. Other examples include the enriched HLA allele DRB1*11:14:01 (CaG AF=0.0012, global reference AF=7.0×10^−5^, P-value=9.7×10^−5^), which has been associated with juvenile idiopathic arthritis (JIA)^30,31^, and HLA-B*51:01 (CaG AF=0.07, global reference AF=0.04, P-value=4.0×10^−13^), which is also associated with adverse drug reaction^32^ and Behçet’s disease^33^. While the enrichment in the first two cases could be explained by drift associated with a single French settler carrying the allele, an increase of 3 percentage points in allele frequency over the same period is not consistent with founder events in Quebec. Rather, it points to a mismatch between the French source population frequency and gnomAD populations of European ancestry (with a higher proportion of UK and Northern Europe ancestry contributions). Indeed, HLA-B51:01 has a frequency of 7% in France^34,35^, and can reach up to 25% in Silk Road nations^35,36^. Thus, variant enrichment in Quebec relative to the rest of North America may not always reflect recent founder events, but rather differences in ancestral composition.

### Variants of clinical interest in *SPG7*

Variants of clinical interest (VCI) involved in disease are typically classified as pathogenic or likely pathogenic based on ACMG guidelines^37^. Knowledge of VCI in a gene contributes to the ability to confirm diagnoses in affected individuals, assess the prevalence of the associated condition, and estimate the population carrier rates for autosomal recessive conditions. We used CaG WGS to compare different approaches to study VCI in the gene *SPG7* (MIM 602783). Pathogenic variants in *SPG7* are associated with an autosomal recessive form of hereditary spastic paraplegia (HSP, MIM 607259) that is characterized by adult-onset progressive bilateral leg weakness and spasticity, sensory neuropathy and in some cases ataxia and dysarthria^38^. Diagnosis is based on typical clinical findings and identification of biallelic pathogenic variants in *SPG7*. HSP due to *SPG7* variants is a frequent cause of spastic ataxia in French Canadians (FC)^39^.

First, we retrieved two well-documented pathogenic *SPG7* variants — c.1529C>T (p.Ala510Val) and c.988-1G>A — in the CaG WGS data (**Fig. 2** and **Supplementary Table 8**)^40^. The c.1529C>T variant was found at a similar frequency in CaG WGS (0.0051) relative to gnomAD NFE (0.0058), whereas c.988-1G>A was significantly enriched (three carriers; odds ratio (OR)=36.6; P-value=8.3×10^−4^). Second, we identified all *SPG7* variants in the CaG WGS data with ClinVar pathogenicity interpretation and found six with previous evidence, of which five passed manual curation for pathogenicity evidence based on ACMG criteria (**Fig. 2** and **Supplementary Table 8**)^37^, including the two variants described above. Third, starting with 2,054 potential pathogenic variants in *SPG7* (including putative loss-of-function and splice site variants), we prioritized nine variants including the six variants identified in the first and second approaches. Amongst the additional three variants, the missense c.2293G>A (p.Asp765Asn) variant was significantly enriched in the CaG cohort (OR=21.9; P-value=0.0019). A new splice variant was also significantly enriched: c.376+1G>T (OR=11.2, P-value=0.032). This shows that putative founder variants identified in studies of affected individuals may in fact be as frequent in regional or worldwide populations (e.g. p.Ala510Val), whereas previously unreported variants are likely to be founder variants (e.g. p.Asp765Asn and c.376+1G>T).

**Figure 2.**
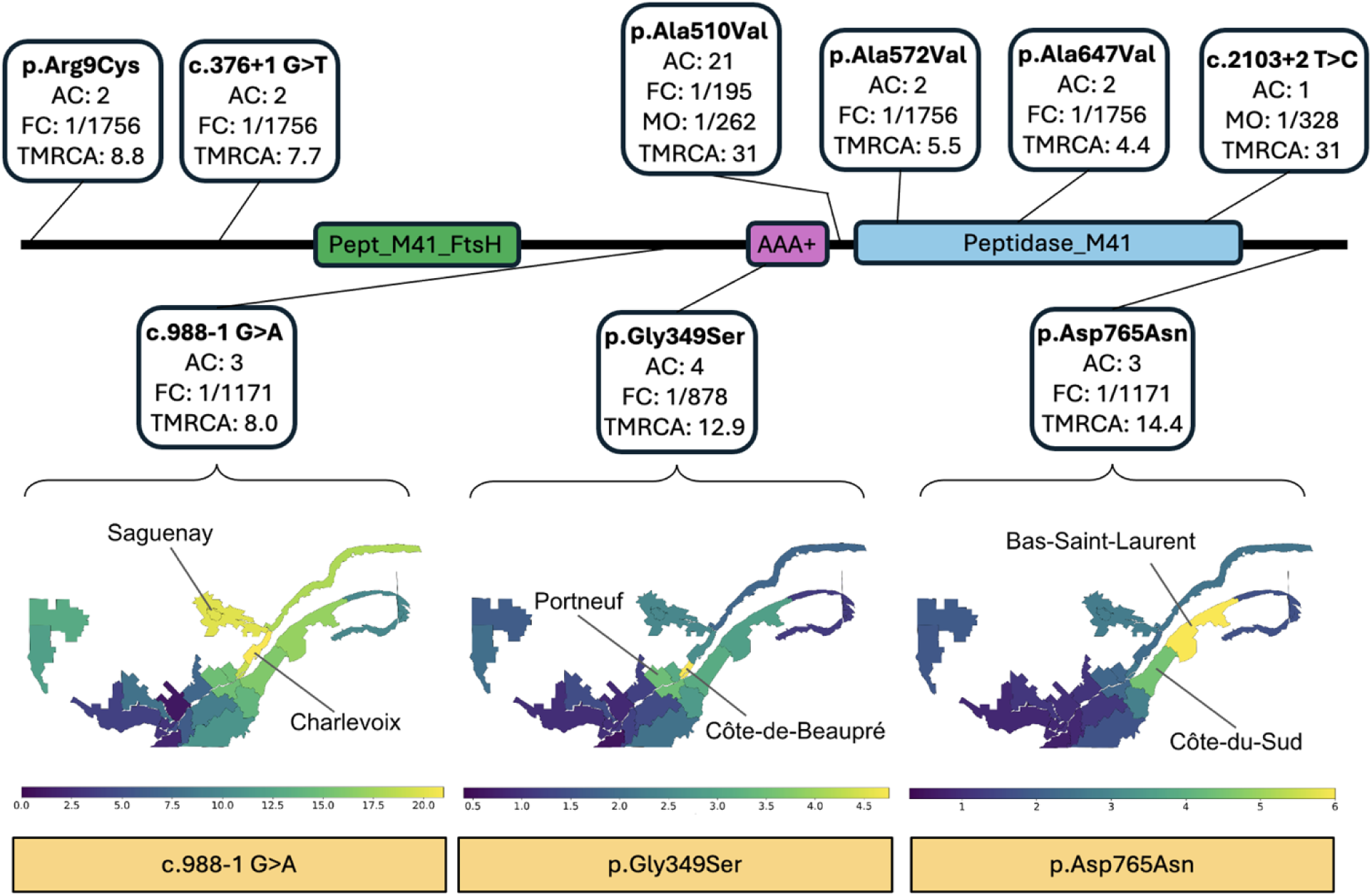
Analysis of rare pathogenic variants in *SPG7*. Structure of the *SPG7* gene, highlighting its main three domains and the location of nine variants found in the CaG cohort. For each variant, amino acid/nucleotide change, allele count (AC), frequency in Quebec residents of French-Canadian ancestry (QFC), and estimated mutation age (TMRCA) are displayed. The heatmaps show the carrier frequency of three *SPG7* variants computed with ISGen in 24 historical regions of Quebec. The scale is given in number of carriers per 1000 individuals. Each variant has a different frequency distribution across Quebec. Highest frequency regions for each variant are highlighted: C.988-1G>A (Saguenay and Charlevoix), p.Gly349Ser (Côte-de-Beaupré and Portneuf) and p.Asp765Asn (Bas-Saint-Laurent and Côte-du-Sud).

Ancestral recombination graph (ARG) reconstruction^41^ of eight of the nine *SPG7* variants described above suggested common ancestry within the last 15 generations, consistent with the introduction of each of these alleles by a single individual in early New France. The exception was c.1529C>T (p.Ala510Val), with a time to most recent common ancestor (TMRCA) of 96 generations (**Fig. 2** and **Supplementary Table 9**), suggesting multiple introductions, consistent with the high gnomAD NFE frequency mentioned above. We used ARG-based imputation to identify likely carriers of each variant among the genotyped CaG individuals, and selected three *SPG7* variants (c.988-1G>A, c.1045G>A, and c.2293G>A) with over 40 carriers linked to the BALSAC genealogical record to perform genealogical analysis using ISgen^42,43^. We identified the same founder family as the most likely to have introduced both c.988-1G>A and c.1045G>A. This family contains two couples who were married in Perche, France in the 1600s. We estimated the allele frequency for the three *SPG7* variants for 23 historic regions of Quebec (**Fig. 2**). Despite a common inferred founder family, results show uneven distribution across regions, likely a result of regional founder events: for c.988-1G>A, we identified Côte-de-Beaupré (carrier frequency 1/210) and Portneuf (1/288) as the regions with highest estimated carrier frequency. For variant c.1045G>A, we estimated highest frequencies in Charlevoix (1/188) and Saguenay-Lac-St-Jean (1/197). Finally, variant c.2293G>A showed higher carrier frequency in Bas-Saint-Laurent (1/167) and Côte-du-Sud (1/225). Thus, founder variants in QFC are strongly affected by regional founder events.

In summary, the analysis of population-based sequence data from individual genes, together with unique historical migration records and genealogies in QFC, can help systematically clarify the prevalence of known and putative novel disease-causing variants and their fine-scale distribution in the population.

### Haplotype reference panel and genotype imputation

Genotype imputation in populations with unique histories benefits from ancestry-matched reference haplotype panel^44^. We assembled a CaG reference panel using haplotypes from 2,173 WGS CaG participants, which included 15,483 SVs, 307 HLA alleles, and ∼44 million SNVs/indels. We assessed the quality of the reference panel using independent samples of 10 family trios and 141 unrelated individuals from Quebec. We observed the best accuracy of statistical phasing in the CaG panel in QFC individuals [average switch error rate (SER)=0.152% and standard error (SE)=0.005] (**Supplementary Fig. 12-14** and **Supplementary Tables 10-12**). Adding prior phasing information from the ∼2 times larger 1000 Genome Project and Human Genome Diversity Project’s combined panel (N=3,941)^45^ to the statistical phasing model provided only minimal gain to phasing accuracy for common variants in QFC individuals (reference panel alternate allele count >200, the average pairwise SER was reduced by 0.003-0.010%), suggesting that the CaG panel captures the majority of QFC-specific haplotypes.

Validation was performed by down-sampling WGS data to genotyping array position from individuals not used in the reference panel (Methods, **Supplementary Fig. 15**). In regions that are not challenging to sequence^46^, the CaG reference panel enabled the imputation of 179,840 (SE=352) SNVs/indels per individual that were missed by the multi-ancestry TOPMed panel (N=133,597) while being present in the original WGS data (**Supplementary Fig. 15A-B**). These had average concordance of 99.33% (SE=0.04%). As expected, variants enriched in CaG were more likely to be well-imputed with the CaG but not with the TOPMed panel: variants with 8-fold enrichment in CaG relative to gnomAD NFE (and with AF<5% in gnomAD NFE), were 3.51-fold more likely to be present in the original WGS, imputed by CaG, but missing in TOPMed-imputed data relative to all variants with frequency below 0.05 in gnomAD NFE (**Supplementary Fig. 15C**). On average, 31,410 (SE=1,635) SNVs/indels per individual were missed by both panels (**Supplementary Fig. 15A**). These variants were also more likely to be enriched in CaG relative to gnomAD NFE (**Supplementary Fig. 15C**, up to 1.66-fold), and to be at low gnomAD allele frequency (**Supplementary Fig. 15D)**, suggesting that allele counts for many variants in CaG were not quite high enough to impute correctly, and that even greater number of QFC reference haplotypes is needed to capture all rare alleles that are more frequent in QFC. We also confirmed the robustness of the CaG panel in imputing SVs and HLA alleles. The concordance between SVs identified using WGS data and imputed values was between 96.3% and 98.8%, depending on the SV type and imputation quality thresholds. Similarly, 96.2-99.1% of imputed HLA alleles were concordant with typed HLA alleles depending on their resolution and imputation quality thresholds (**Methods** and **Supplementary Notes)**.

### Genome-wide association studies

We performed GWAS on 42 continuous traits (**Supplementary Table 13**) using genotype dosages from imputation with the CaG panel, with the TOPMed panel, and with the meta-imputation of these two reference panels (**Methods**). Despite containing ∼75 times fewer reference haplotypes than the multi-ancestry TOPMed panel, the CaG panel sampled from the study population allowed for the identification of more independent genome-wide significant loci when analyzing CaG participants of European genetic ancestry (378 loci with CaG vs. 356 with TOPMed, N=27,037) or all participants (482 loci with CaG vs. 452 with TOPMed, N=29,334) (**Supplementary Table 14**). The meta-imputation approach showed the largest number of genome-wide statistically significant loci, especially in the multi-ancestry case (379 in European-ancestry and 515 in all ancestries).

The overlap in identified GWAS loci between different imputation approaches was high but not complete (**Fig. 3** and **Supplementary Fig. 16**). Among 58 additional loci identified using only the CaG panel, 23 (∼40%) had a lead variant absent in the TOPMed panel (**Fig. 3**). In contrast, among 33 additional loci identified when using only the TOPMed panel, only six (∼18%) lead variants were absent from the CaG panel (**Fig. 3**). Thus, the increased variant coverage when using local reference panels seems critical to uncover additional association signals, while larger reference panels may help improve the imputation accuracy of more common variants present in both datasets. In general, the TOPMed panel resulted in better imputation quality scores than CaG for variants with AF < 0.01, while CaG showed slightly better scores for variants with AF ≥ 0.01 (**Supplementary Fig. 17**). The imputation quality was better for lead variants at significant GWAS loci in the CaG-imputed than in the TOPMed-imputed data, except for loci shared between TOPMed and meta-imputation results only (subset D, **Fig. 3B**). The largest improvement in imputation quality (median pairwise difference of 0.034) was found at loci shared only between CaG and meta-imputation results (subset B). This improvement was higher than expected for the same number of variants with similar AF across the entire genome (two-tailed P-value=0.009 after 1,000,000 permutations). Thus, local reference panels may provide additional GWAS signals through improved imputation of variants that are already available in global panels (**Fig. 3** and **Supplementary Fig. 16**).

**Figure 3.**
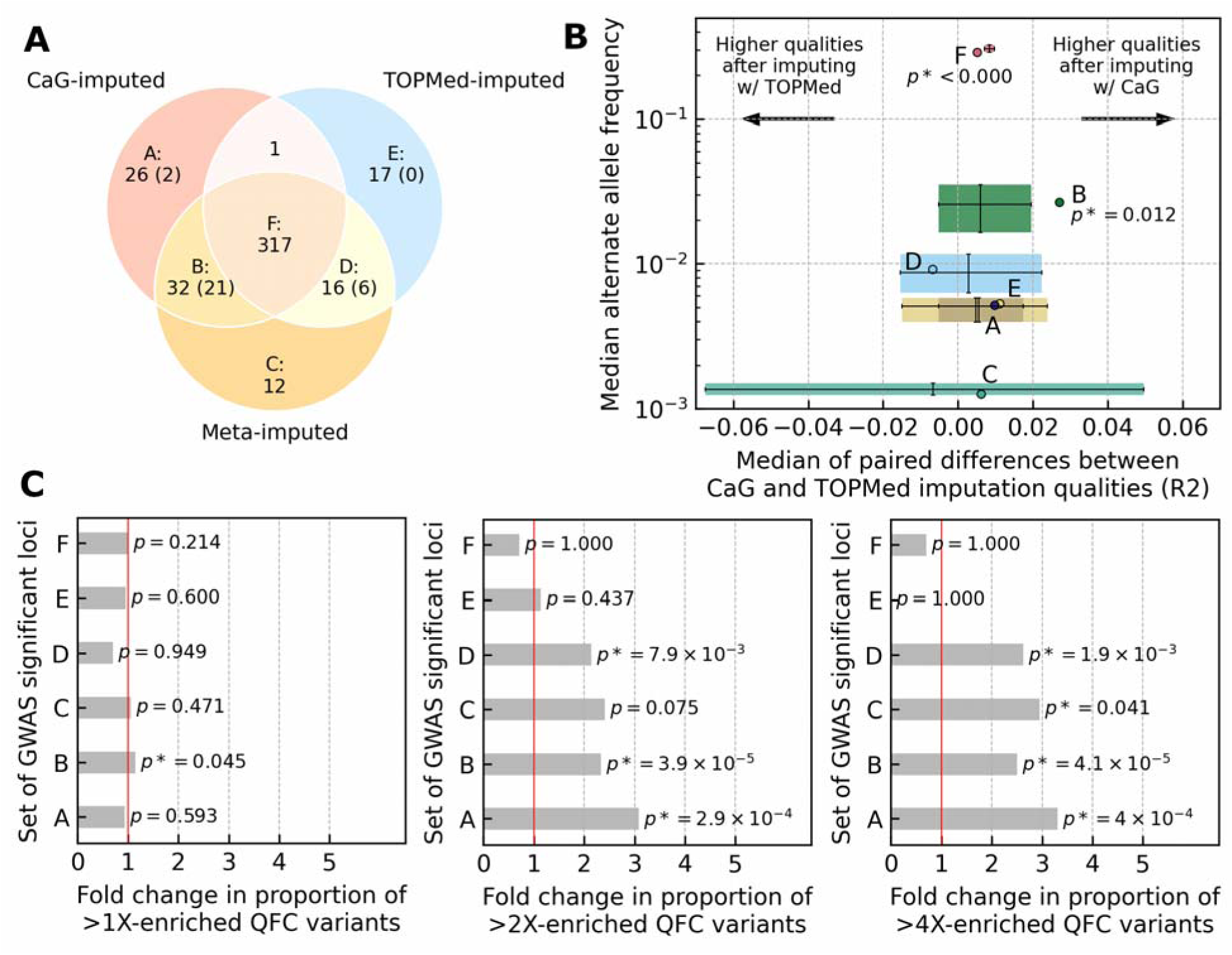
Genome-wide statistically significant loci identified using different genotype imputation approaches in individuals of genetic European ancestry. The statistically significant independent locus was defined as the ±500-kb region around the variant with the lowest statistically significant P-value, referred to as the lead variant. Any overlapping loci for the same trait were merged into a single locus, keeping one lead variant with the lowest P-value. Thus, by definition, each locus had only one lead variant. (**A**) The overlap between genome-wide statistically significant loci using three imputation approaches. Letters A, B, C, D, E, and F label the corresponding loci subsets. The number below the label shows the number of loci within each subset. The number in brackets corresponds to the number of lead variants not found in the alternative reference panel. (**B**) The X-axis shows the median of paired differences between imputation qualities at lead variants in CaG and TOPMed imputation results. The Y axis shows the median alternate allele frequency (AF) of lead variants based on CaG imputation results. The horizontal and vertical error bars show 95% confidence intervals after 1,000,000 permutations stratified by AF. Only statistically significant (significance threshold 0.025) two-tailed permutation P-values for the median of paired differences in imputation qualities are displayed below the subset labels to reduce image cluttering. (**C**) Each panel shows a fold change in the proportion of lead variants inside different subsets of significant loci, from left to right: lead variants for which alternate allele frequency (AF) in QFC individuals exceeds AF in gnomAD NFE, lead variants for which AF in QFC individuals is 2 times higher compared to gnomAD NFE, lead variants for which AF in QFC individuals is 4 times higher compared to gnomAD NFE. The P-values were computed as the proportion of 1,000,000 permuted samples that exceeded the observed fold change with a statistical significance threshold of 0.05.

To facilitate access to the CaG GWAS results and enable replication studies, we released the single-variant association results into a publicly available PheWeb browser (https://cerc-genomic-medicine.ca/pheweb/cartagene/). We also summarize the GWAS results for the imputed SVs and HLA alleles in the **Supplementary Notes**, **Supplementary Fig. 18** and **Supplementary Tables 15-16**. In this section, we emphasize two noteworthy examples that demonstrate the advantages of using the CaG imputation panel to perform GWAS in the Quebec population. First, a one base-pair deletion (rs796144540) inside an intron of the insulin receptor gene *INSR* was associated with increased thyroid-stimulating hormone (TSH) levels (MAF=0.42, P-value=3.3×10^−8^) (**Supplementary Fig. 19**). This variant was absent from the TOPMed panel, presumably because it was missed during sequencing or removed at QC, but well-imputed with the CaG panel (INFO=0.96). Conditional analysis controlling for rs796144540 identified a second independent rare *INSR* intronic variant associated with TSH levels (rs8107575, INFO=0.85, MAF=0.001, P-value=8.5×10^−6^). Thyroid abnormalities were previously reported in patients with mutations in *INSR*^47^. Second, we identified an association between basophil counts and a rare intergenic variant (rs867418280, INFO=0.77, MAF=0.0032, P-value=3.7×10^−8^) that was well-imputed with the CaG haplotypes but absent from the TOPMed imputation panel (**Supplementary Fig. 20**).

We found that several sentinel variants that were well-imputed with the CaG panel or the meta-imputation were more frequent in QFC compared to gnomAD NFE (Sets A, B and C in **Fig. 3**). This observation motivated a secondary analysis in which we focused on associations identified between the same 42 quantitative traits and well-imputed, high-impact variants that are enriched at least four-fold in QFC relative to gnomAD NFE (**Supplementary Fig. 21**). For these analyses, we considered both enriched high-impact coding (see above) and non-coding variants (annotated as conserved or epigenetically active by FAVOR, **Methods**). We selected variant-phenotype pairs with a P-value <1×10^−5^ in CaG and attempted their replication in the Canadian Longitudinal Study on Aging (CLSA, N=23,752), which includes 4,602 individuals recruited in Quebec (**Methods**). We were limited to variants and traits also assessed in the CLSA; in total, we replicated 6/15 (40%) variant-phenotype pairs at a nominal P-value of <0.05 with the same direction of effect (**Table 1**; an additional pair was replicated in the UK Biobank, see below). The frequency of the tested variants and the limited number of Quebec residents in CLSA to match the CaG analyses may explain this low replication rate.

**Table 1.** Selected variants that are enriched in CaG QFC participants and associated with clinically relevant quantitative traits. When available, we attempted to replicate associations in the European genetic ancestry subset of the Canadian Longitudinal Study on Aging (CLSA) or the UK Biobank (for the *PDZK1* variant). Coordinates are on build GRCh38, and the direction of the effect is for the alternate (ALT) allele. In bold are variants that replicate at nominal significance (P-value <0.05). FEV1 = forced expiratory volume; HDL-C = high density lipoprotein cholesterol; RBC = red blood cell count; TRIG = triglycerides; TSH = thyroid stimulating hormone

| Trait | Variant ID | CHR_POS_REF_ALT | CaG |  |  | CLSA |  |  | gnomAD<br>NFE (v4.1.0)<br>FREQ_ALT | Note |
| --- | --- | --- | --- | --- | --- | --- | --- | --- | --- | --- |
|  |  |  | FREQ_ALT | BETA (SE) | P-value | FREQ_ALT<br>(FREQ_ALT<br>Quebec<br>resident<br>subset) | BETA (SE) | P-value |  |  |
| FEV1 | rs184696760 | chr5_964735_G_A | 0.0015 | -0.594<br>(0.112) | 1.3x10<sup>-7</sup> | 2.6x 10<sup>-4</sup><br>(0.0011) | 0.172<br>(0.209) | 0.41 | 1.5x10<sup>-4</sup> | APC_Conservation<br>(0.02);<br>APC_Epigenetics<br>(21.5) |
| HDL-C | rs118204060 | chr8_19954279_C_T | 0.0022 | -0.752<br>(0.085) | 9.5x10<sup>-19</sup> | Variant not available |  |  | 3.1x10<sup>-5</sup> | LPL p.Pro207Leu |
| HDL-C | **rs548046676** | chr9_108086996_T_G | 0.0028 | -0.421<br>(0.073) | 8.5x10<sup>-9</sup> | 0.00056<br>(0.0015) | -0.503<br>(0.170) | 0.0031 | 0.00059 | APC_Conservation<br>(21.4);<br>APC_Epigenetics<br>(5.2) |
| HDL-C | rs905396624 | chr9_110152916_C_T | 0.0022 | -0.558<br>(0.085) | 5.7x10<sup>-11</sup> | 0.00014<br>(0.00068) | -0.020<br>(0.334) | 0.95 | 2.9x10<sup>-5</sup> | APC_Conservation<br>(8.8);<br>APC_Epigenetics<br>(20.6) |
| HDL-C | **rs1282267586** | chr16_58243922_A_G | 0.0013 | 0.513<br>(0.108) | 1.9x10<sup>-6</sup> | 0.00039<br>(0.0016) | 0.559<br>(0.205) | 0.0064 | 4.4x10<sup>-5</sup> | APC_Conservation<br>(2.9);<br>APC_Epigenetics<br>(20.3) |
| HDL-C | rs1022447460 | chr16_66830714_G_A | 0.0020 | 0.418<br>(0.088) | 2.3x10<sup>-6</sup> | 0.00070<br>(0.0029) | 0.299<br>(0.154) | 0.052 | 7.4x10<sup>-5</sup> | APC_Conservation<br>(10.2);<br>APC_Epigenetics<br>(25.5) |
| HDL-C | rs781563144 | chr16_68732415_C_A | 0.0018 | 0.424<br>(0.089) | 1.7x10<sup>-6</sup> | 0.00066<br>(0.0028) | 0.298<br>(0.159) | 0.060 | 5.9x10<sup>-5</sup> | APC_Conservation<br>(11.6);<br>APC_Epigenetics<br>(21.2) |
| Platelet | **rs906008790** | chr12_53974862_G_A | 0.0036 | -0.382<br>(0.086) | $9.0 \times 10^{-6}$ | 0.00054<br>(0.0025) | -0.570<br>(0.191) | 0.0028 | $7.9 \times 10^{-5}$ | APC_Conservation<br>(6.2);<br>APC_Epigenetics<br>(22.9) |
| Platelet | **rs1317562039** | chr12_54295902_G_C | 0.0037 | -0.402<br>(0.084) | $1.8 \times 10^{-6}$ | 0.00059<br>(0.0028) | -0.473<br>(0.184) | 0.010 | NA | APC_Conservation<br>(23.2);<br>APC_Epigenetics<br>(18.9) |
| RBC | **rs755847829** | chr16_636735_G_C | 0.0014 | 0.780<br>(0.130) | $1.7 \times 10^{-9}$ | 0.00054<br>(0.0027) | 0.770<br>(0.180) | $1.40 \times 10^{-5}$ | 0.00013 | APC_Conservation<br>(0.001);<br>APC_Epigenetics<br>(24.2) |
| TRIG | rs118204060 | chr8_19954279_C_T | 0.0021 | 0.610<br>(0.095) | $1.4 \times 10^{-10}$ | Variant not available | | | $2.9 \times 10^{-5}$ | LPL p.Pro207Leu |
| TRIG | rs991735459 | chr11_117979750_T_C | 0.0015 | 0.532<br>(0.107) | $6.3 \times 10^{-7}$ | 0.00051<br>(0.0019) | 0.166<br>(0.197) | 0.40 | 0.00012 | APC_Conservation<br>(0.1);<br>APC_Epigenetics<br>(21.9) |
| TSH | rs780939824 | chr5_168263763_C_A | 0.0023 | 0.417<br>(0.092) | $6.0 \times 10^{-6}$ | 0.00021<br>(0.00018) | -0.175<br>(0.319) | 0.57 | 0.00035 | APC_Conservation<br>(20.3);<br>APC_Epigenetics<br>(5.2) |
| TSH | **rs121908863** | chr14_81092547_C_G | 0.0015 | 0.688<br>(0.111) | $5.2 \times 10^{-10}$ | 0.00052<br>(0.0016) | 0.684<br>(0.196) | 0.00048 | 0.00029 | TSHR<br>p.Pro162Thr |
| Uric acid | **rs191362962** | chr1_145688000_G_A | 0.0014 | -0.673<br>(0.137) | $8.4 \times 10^{-7}$ | Phenotype not available | | | 0.00018 | PDZK1 p.Arg9ter;<br>associated with<br>lower uric acid<br>levels in the UKBB<br>($P=2.3 \times 10^{-13}$) |

The list of replicated variant-phenotype pairs includes the well-known *LPL* founder p.Pro207Leu variant, associated with increased HDL-cholesterol (HDL-C) and triglyceride levels, which accounts for ∼72% of all pathogenic *LPL* alleles found in the QFC population (**Table 1**)^48^. The Leu allele at this variant is 48 times more frequent in QFC than in gnomAD NFE. We also found a rare nonsense variant (p.Arg8ter, MAF=0.1%) in the *PDZK1* gene that associates with lower uric acid levels; the variant is 10 times more frequent in QFC than in gnomAD NFE (**Table 1**). Common variants near *PDZK1*, which encodes a urate transport accessory molecule, have been associated with uric acid levels and gout risk^49,50^. The variant is missing in the CLSA, but in the UK Biobank, the coding *PDZK1* p.Arg8ter variant (MAF=0.02%) is associated with lower uric acid levels (P-value=2.3×10^−13^) and protection against gout (OR=0.32, P-value=0.024)^51^. We identified novel associations between HDL-C and two rare non-coding variants (**Table 1**). We also found two non-coding variants associated with platelet count, although these two variants likely represent a single association signal as they are in strong LD (*r*^2^=0.91; D’=0.97) (**Table 1**). Finally, we found an association between a rare coding variant (p.Pro162Thr) in the TSH receptor gene (*TSHR*) and TSH levels (**Table 1**). This *TSHR* variant is 5-times more frequent in QFC than in gnomAD NFE participants and has previously been found in patients with congenital hypothyroidism and nonautoimmune hyperthyrotropinemia^52,53^.

### Expression quantitative trait loci analyses

To generate mechanistic hypotheses that may explain our GWAS results, we performed eQTL analyses with 504 individuals for whom WGS and whole-blood RNA-sequencing (RNAseq) data was available^54^. Focusing on variants with a MAF >1%, we identified 2,223,590 significant associations (eSNPs) implicating 12,774 genes (eGenes) at a false discovery rate (FDR) <5% (**Methods**). We observed a strong concordance between our CaG results and the significant whole-blood eQTL results from GTEx v8 (N_eQTL_=1,523,724, Spearman’s ρ=0.82, P-value<2.2×10^−16^) (**Supplementary Fig. 22A**). Many eQTL association results (42.5%) were more significant in CaG than in GTEx despite a similar sample size, potentially due to differences in the RNAseq experimental protocol or design, the genetic ancestry of the populations analyzed or their environmental exposure (**Supplementary Fig. 22B**)^54^. All CaG eQTL results are available through a publicly available browser (https://eqtlbrowser-cag.mhi-omics.org/).

## DISCUSSION

Many studies have previously leveraged the founder effect in the QFC population to identify disease-causing variants. These studies often had a small sample size and were limited to specific disease cohorts or families. Thus, there have been no efforts to characterize at scale genetic variation in the multi-ancestry and ethnically diverse population of Quebec. To address this knowledge gap, we profiled and analyzed genetic information from 29,337 participants from the population-based cohort CaG. Despite the wealth of genetic information already available for several populations worldwide, including individuals of genetic European ancestry, there is a clear added value to continue to characterize the genetic variation in populations with unique history and demography. For instance, our analyses of the CaG genetic data allowed us to: (1) gain insights into the genetic diversity and demography of modern Quebec, (2) prioritize clinically relevant variants in *SPG7* and estimate their AF in different regions of Quebec, and (3) develop better genotype imputation tools that have enabled the discovery of genetic associations missed using global haplotype reference panels.

While it is currently the largest population-based cohort in Quebec, and represents a range of ancestral backgrounds, CaG is not completely representative of the province’s population. On average, CaG participants have higher education attainment and income than the overall population. Furthermore, because recruitment took place in six cities, several geographical regions of Quebec such as the Northwestern Abitibi-Temiscamingue region are not well captured in the CaG dataset. Indigenous residents are also not well represented in CaG. This will require continued efforts and engagement with these communities to support equitable access to the benefits of advances in human genetics and their impacts in medicine. Finally, we preferentially selected participants of Haitian and Moroccan descent in the WGS study, as they help capture variation relatively common in Quebec but poorly characterized internationally.

The new datasets will also be relevant to many individuals outside Quebec. Large communities of individuals with FC ancestry live throughout Canada and in the Northeastern United States. The WGS data for 163 QHA and 132 QMO CaG participants forms one of the largest resources available for these ancestry groups with large populations in Haiti, Morocco, and in diasporas around the world. Integrating the CaG genetic data with other sequencing projects will facilitate the development of genetic resources that are tailored for the Haitian and Moroccan populations^55^. While such data integration presents ethical and legal challenges, summary level data sharing via our GWAS and allele frequency browsers provides a practical first step.

## METHODS

### CARTaGENE

The CARTaGENE (CaG) Cohort recruited 43,032 participants (40-69 years old at recruitment) across five different regions in the province of Québec, Canada. Participants have each provided informed consent. The cohort has been described elsewhere^8^ and **Supplementary Table 1** provides a summary of its demographics. 29,337 participants provided biospecimens, including whole blood from which we extracted DNA. CaG is approved by the Ethics Committee of the CHU Ste-Justine. A local Institutional Review Board (IRB) for each investigator involved in this study has also approved genetic analyses of the CaG data.

### Genome-wide DNA genotyping

Most CaG participants have been genotyped on a version of the GSA arrays (https://cartagene.qc.ca/files/documents/other/Info_GeneticData3juillet2023.pdf).The genotyping was done separately in five different projects, each time using a slightly different array. We used PLINK 1.9^56^ to merge these five datasets together, and filter to remove duplicated individuals, and duplicated or monomorphic variants. We also filtered indels and variants with either a high missingness (>5%) or Hardy-Weinberg disequilibrium, or both (--hwe 0.000001 midp --geno 0.05).

### Whole-genome DNA sequencing (WGS)

#### Participant selection

We selected 2,184 unrelated participants (based on identity-by-descent sharing, <3^rd^ degree familial relationship) for the WGS phase of this project, of which 2,173 passed quality control. These included 1,878 European-ancestry individuals with four grandparents born in Canada falling within the majority cluster in PCA space (including 1,756 QFC), as well 163 and 132 participants with four grandparents born in Haiti (QHA) and Morocco (QMO), respectively. Other selection criteria included: (1) availability of genome-wide genotyping array data, (2) possibility to re-contact via email for future studies, (3) availability of whole-blood RNA-sequencing data (N=504), and (4) linkage with the BALSAC genealogy database (N=1,074).

#### DNA sequencing and variant calling

DNA samples were sequenced at the Genome Quebec *Centre d’Expertise et de Services* (CES) on Illumina NovaSeq sequencers using PCR-free libraries and a paired-end (2×150 bp) protocol. The mean coverage was 33.4X (range 16.3 to 72.0). We used the Illumina DRAGEN pipeline (v3.9.5) to call single nucleotide variants (SNVs) and short insertion-deletions (indels, <50-bp) at the sample level, and across the complete WGS dataset to create a multi-sample VCF, using default parameters. Before quality control, the dataset included 64,503,029 SNVs and 7,025,566 indels: 27,915,550 singletons, 8,693,233 doubletons, and 3,638,863 multi-allelic variants.

#### WGS data quality-control

We used PLINK 1.9 to perform quality control on the CaG WGS dataset. We excluded variants with >5% data missing, as well as samples with >5% data missing or with extreme heterozygosity (|Fstat| >0.4). We computed HWE statistics for each variant and reported those in a supplementary file available from the CaG data access portal. We used bcftools v1.16 (options “norm -m -both”) to convert multi-allelic variants into multiple bi-allelic variants. Our analysis with the seGMM software to analyze the karyotype of sex chromosomes did not detect sex mismatches^57^. The final dataset included 2,173 individuals and 80,407,530 variants, of which 62,457,187 are bi-allelic SNVs, 6,698,539 are bi-allelic indels, and 11,251,804 are bi-allelic variants (SNVs or INDELs) converted from 3,148,393 multi-allelic variants (**Supplementary Table 2**). The concordance between genotypes from the WGS and genotyping (Illumina GSA arrays) variant call sets for these individuals and calculated from >645,000 variants is >99.87%.

#### Variant annotation

We calculated the allele frequency of each variant in the whole WGS dataset as well as within each of the three ancestral groups and provide this information in supplementary files (on the data access portal) as well as on our allele frequency browser (https://cartagene-bravo.cerc-genomic-medicine.ca/). We annotated variants using the ENSEMBL Variant Effect Predictor (VEP) module^58^ and FAVOR^59^; these annotations are also available as supplementary files through the CaG data access portal. VEP v108 was run locally using the following parameters: “--symbol --pick --check_existing --af_gnomadg --sift b --polyphen b”. High-impact variants (as highlighted by VEP with ‘IMPACT=HIGH’), belong to these categories: frameshift, stop gained and loss, splice donor and acceptor, start loss, and transcript ablation, resulting in 26,874 variants, of which 23% are novel (not found in dbSNP version 154). To this list, we added 980 variants defined as (strictly) pathogenic in the ClinVar database. Thus, the list of high-impact coding variants considered for downstream analyses included 27,361 variants. To prioritize non-coding variants, we used composite scores (aPC ≥20) generated by FAVOR that consider multiple variant annotations based on conservation (aPCcons) or epigenomic (aPCepi) scores^59,60^. From the list of high-impact coding and non-coding variants, we then defined a list of enriched high-impact variants (hereafter referred to as enriched variants) using these two criteria: (1) a minor allele frequency (MAF) in CaG ≥0.1% and (2) a ratio of allele frequency (CaG) / allele frequency (gnomAD) ≥4. From gnomAD, we used allele frequencies from Non-Finnish European-, African-, and Middle Eastern-ancestry populations as reference for the QFC, QHA and QMO participants, respectively. In total, we found 1,375, 2,639, and 2,970 enriched coding variants in QFC, QHA, and QMO participants, respectively. In non-coding regions, we found 7981 variants with high aPCcons and 20,896 variants with high aPCepi that were enriched in QFC.

### Population genetic analyses

#### Principal component (PC) analysis and uniform manifold approximation projection (UMAP)

We generated principal components (PCs) using PLINK. We pruned for linkage disequilibrium using PLINK2 (parameter --indep-pairwise 1000 50 0.1). We generated PCs using PLINK1.9, masking chromosome 6 from 25000000 to 33500000 to filter out HLA due to the long-range LD patterns in this region and using parameters: --maf 0.5 --mind 0.1 --geno 0.1 --hwe 1e-6. To calculate the percentage of variance explained, we used the R implementation of flashpca^61^.

We generated UMAP coordinates using the umap-learn package (v0.5.1) in Python^62^. For the visualization, we ran UMAP on the top 10 PCs using the 15 nearest neighbors and a minimum distance of 0.5 to focus on visual clarity^10,62^. Colors for the PC analysis and UMAP visualizations were based on clusters defined via density clustering as described in Diaz-Papkovich et al.^10^. We used HDBSCAN (v0.8.27) with parameters Ê = 0.3 and a minimum cluster size of 25 points on a 3D UMAP of the top 25 PCs parameterized for the 10 nearest neighbors and a minimum distance of 0.001. Word clouds, created with the Python wordcloud package (https://github.com/amueller/word_cloud), were generated using the country of birth, province of birth (for those born in Canada), selected ethnicity, and collection region variables.

#### Admixture analysis

We used genotype data to estimate ancestry proportions for three populations with WGS data available: European-ancestry with four grandparents born in Canada (including QFC and non-QFC participants), QMO and QHA. To highlight population structure within each country of origin, we selected individuals that reported their four grandparents to be born in the same country. For each of the populations of interest, we used KING with option --kinship to identify kinship relationships between individuals. We filtered out related individuals to avoid relations closer to 3^rd^-degree (Kinship coefficient > 0.0442) using PLINK1.9 flag –king-cutoff. We found four pairs of related individuals in the QHA dataset: 3 pairs at 3rd-degree and 1 pair at 1st-degree. One individual per pair was removed. No individuals were removed from the QMO dataset. The final population sets were composed of 267 QHA, 201 QMO, and 500 randomly sampled individuals with four grandparents born in Canada (which includes, but is not limited to, QFC).

To study the population structure of each group of interest, we built three datasets with populations based on similar geographical and historical background. For CaG-based ancestry references, we selected participants with four grandparents born in the same country. The reference dataset for QMO included CaG participants with ancestry from North and Sub-Saharan Africa, the Middle East, and Western Europe. The reference dataset for QHA included CaG ancestry from Sub-Saharan Africa and Western Europe, along with four populations from the 1000 Genomes Project Phase 3 (YRI, ACB, CEU, and CHB). For the continental ancestry estimation in individuals with grandparents in Canada, our dataset included French, British, Latin American and Sub-Saharan African ancestries from CaG.

We used the ADMIXTURE algorithm in unsupervised mode to estimate ancestry proportions from the merged genotype data^63^. We filtered genotypes for LD using PLINK1.9 with parameter --indep-pairwise 50 10 0.5. This left 421,356, 403,640, and 204,003 variants after pruning for the Canadian (with four grandparents born in Canada), Moroccan (QMO), and Haitian (QHA) datasets, respectively. Each of the three pruned datasets were run independently for admixture estimation. We performed three independent runs of admixture at each value of K for K=3-7 (QHA and QMO), and K=2-5 (CA). Optimal K was determined by computing the cross-validation error in each run, option –cv in ADMIXTURE. Visualization of ancestry components across multiple runs and Ks was done with PONG^64^.

#### IBD detection and clustering

Shared genetic variants from the five genotyping arrays in CaG were used to detect identity-by-descent (IBD) segments. Variants were phased using Beagle 5.4 (beagle.22Jul22.46e.jar) and the plink.chrCHR.GRCh38.map genetic map from the HapMap project. (http://faculty.washington.edu/browning/beagle/beagle.html). We then filtered the merged file using plink --mac 1 --hwe 0.000001 --max-missing 0.95, resulting in a dataset of 29,330 individuals and 356,441 variants. IBD detection was performed using the HapIBD tool^65^ with default parameters. IBD segments overlapping genome gap regions (short arm, heterochromatin, telomere, contig, scaffold and centromere) were removed. We utilized Louvain clustering methodology, implemented through the igraph R package, to partition individuals into distinct groups based on the total IBD sharing between pairs of samples. Each individual was represented as a node in the network, with edges denoting the extent of IBD sharing. Following three iterations of Louvain clustering, a total of 56 clusters were identified. To mitigate biases introduced by small population sizes or excessive variation in population composition, we computed F_st_ values using plink2 between clusters containing more than 10 samples. Notably, clusters 48 and 54 were excluded from subsequent analyses as they had less than 10 individuals, resulting in a final dataset comprising 29,209 samples. Subsequently, we merged clusters exhibiting F_st_ values below 0.0005, yielding a refined set of 21 clusters of varying sizes, as detailed in the **Supplementary Notes**.

### Identification of variants of clinical interest in *SPG7*

Using CaG WGS sequencing data, we extracted all variants found in any transcripts of the *SPG7* gene. We selected variants classified as Pathogenic (P), Likely Pathogenic (LP), or with conflicting interpretations of pathogenicity according to ClinVar (accession date: March 2024), and high confidence putative loss-of-function (pLoF) variants. Variants were filtered for MAF ≤1% in gnomAD (version v3.1.2), CADD ≥15^66^ and for putative splice variants with a Splice AI score ≥0.8^67^. Then, we performed subsequent manual curation of pathogenicity evidence in databases such as ClinVar and LOVD as well as literature review. Variants were classified using the ACMG 2015 revised guidelines using five distinct categories (benign, likely benign, uncertain significance, LP and P)^37^. Variants with evidence supporting LP or P were considered for further analysis. To calculate enrichment, we estimated the allele frequency for each variant as the count of alternate alleles divided by the total number of alleles as previously described^68^. The allele frequencies observed were compared to those observed in gnomAD, version v3.1.2.

### ARG-based imputation and regional frequency estimation

We used phased genotype data (659,029 variants) to reconstruct the ancestral recombination graph (ARG) for all the individuals in the CaG cohort using ARGneedle^41^. VCF files were pre-processed to keep only biallelic SNPs using plink2 –max-alleles 2 and --snps-only flags, for a total of 566,073 variants. Moreover, genetic map files for the GRCh38 reference genome were obtained in plink format, and interpolation was performed to include all SNPs from the VCF files in the map using an inhouse R script (https://github.com/almejiaga/ARG_needle). Variants with a distance in cM lower than 0.00001 were excluded from the analysis due to ARG-needle software requirements. Finally, VCF files were converted to the Oxford HAP format using plink2. At the end, a total of 548,663 variants were retained for subsequent analyses. The ARG was reconstructed using the ARG-needle software in genotype mode with default parameters for chromosomes 1-22. After, ARG files were converted to tree sequences in the tskit format using the arg2tskit ARG-needle function. We analyzed nine pathogenic mutations in *SPG7*, and we checked in the ARG if carriers share a recent common ancestor among them. For that purpose, we used TSKIT (https://github.com/tskit-dev/tskit) to visualize the ARGs and to verify if the patterns of coalescence between the carriers are consistent with a founder effect. We computed time to the most recent common ancestor of carriers of a mutation using the tmrca function in TSKIT. For imputation, we assumed that all individuals below this MRCA carry the mutation. For mutations with more than 40 carriers, we used the BALSAC genealogy to perform allele frequency estimation within the 24 historic regions using the ISGen software^42^. For this analysis, 100,000 climbing simulations from carriers to founders were performed and 100,000 trees were used for allele frequency estimation; confidence intervals were calculated using bootstrapping with 100,000 iterations.

### Structural variants (SVs)

#### SV calling and genotyping

We detected SVs using Manta (implemented in DRAGEN)^69^ and DELLY 1.1.3^70^ in each participant with WGS data available. We filtered the two SV sets with bcftools 1.11 to keep only variants that had sufficient read support (tag ‘PASS’ in the ‘FILTER’ column of the vcf files)^71^. We used SURVIVOR 1.0.7 to merge the Manta and DELLY filtered SV sets (minimum variant length 50-bp and breakpoint variant overlap 100-bp)^72^. We then used muCNV 1.0.0 (from the working branch of the GitHub repository) to genotype SVs and generate a harmonized multi-samples vcf file: (1) we created a binary interval file of candidate SVs, (2) we generated pileups from each individual’s CRAM file using our candidate list of SVs, and (3) we genotyped each chromosome separately (without merging pileups). We performed an additional filtering step to identify SV pairs that we considered to be the same SV because of the proximity of their start and end coordinates. Two SVs were considered a pair if both their start and end positions were within a window of ≤100-bp, even after the intersection and merging steps done with SURVIVOR. We identified 465 SV pairs with a home-made python script and made sure that both SVs in a pair were of the same type. Next, we compared the allele counts of the two SVs in a pair. For the pairs with the exact same allele counts, we filtered out the SV with the smallest length. For the remaining pairs, we selected those with a percentage of difference between the allele counts and a percentage of difference between the allele frequencies lower than 1% and filtered out the SV with the lowest allele count, assuming that they were the same SVs. We filtered out 301 SVs in total.

#### Comparison of SV allele frequencies with gnomAD-SV

We used the gnomAD SV bed files and vcf files version 2.1 for GRCh37^22^. We lifted over the bed file from GRCh37 to GRCh38 using CrossMap^73^. We split the lifted-over gnomAD bed file into deletions and duplications, and the final SV set with the bcftools filter command. We used bedtools version 2.30.0 intersect (minimum of 50% reciprocal overlap required) to keep the SVs that were common between the gnomAD bedfile and the CaG final SV set vcf file, processing deletions and duplications separately.

### HLA genotyping

We extracted the extended HLA region (chr6:25Mb–35Mb) and performed HLA typing for 11 classical class I and class II HLA genes (HLA-A, HLA-B, -C, -DPA1, -DPB1, -DQA1, -DQB1, - DRB1, -E, -F, and -G) to G-group resolution using HLA*LA^25^. After filtering on allele inference quality (Q_HLA*LA_=1.0) and proportion of bases with >10x depth at each HLA gene locus, and removing samples with >3 HLA alleles missing, 43,150 HLA alleles were retrieved from 2,088 samples. HLA allele frequencies were calculated in the entire WGS cohort and within each ancestral group. Differences in HLA allele frequencies were calculated via Fisher’s exact test, with P-value threshold adjusted for multiple testing via Bonferroni correction, according to the number of alleles compared per group. Global and population-specific HLA allele frequencies were compared to the largest publicly available multi-ancestry HLA reference panel (N= 21,546)^27^ and to low-resolution HLA types previously reported in the Héma-Québec donor biobank^26^ for the overall WGS cohort and stratified by Quebec region of origin.

### Imputation reference panel and genotype imputation

#### Preparing independent samples for statistical phasing benchmarking

We used 40 samples from 10 families (mother, father, twins) collected from the Quebec Study of Newborn Twins^74^. The genomes of samples were sequenced with the Illumina HiSeq 2000 platform at >30X average depth of coverage. We remapped the paired-end reads from the original mapping on the GRCh37 (hg19) to GRCh38 (hg38) human genome assembly using bwa-mem (v0.7.17-r1188)^75^. We performed joint-variant calling using GATK (v4.3) and following GATK’s Best Practices^76^. Using the pedigree information, we performed genotype refinement (GATK’s CalculateGenotypePosteriors, VariantFiltration, and VariantAnnotator tools) to mark *de novo* variants and Mendel law violations. We used only genotypes that pass all GATK’s quality filters in the downstream analyses. We performed a pedigree-based phasing of the genotypes in children using the makeScaffold utility in SHAPEIT4^77^. Using our IBD analyses, we confirmed that these samples cluster together with QFC from CaG. We also randomly selected 50 trios (father, mother, child) from the 1000 Genomes Project and Human Genome Diversity Project (1GP+HGDP) (N=4,904) dataset. The 1GP+HGDP dataset was phased using the pedigree information as part of the gnomAD v3 project^45^. The selected trios included an equal number (N=10) of African (AFR), Admixed American (AMR), East Asian (EAS), Central South Asian (CSA), and European (EUR) genetic ancestries.

#### Preparing independent samples for genotype imputation benchmarking

We used 141 unrelated samples collected in the laboratory of Dr. Guy Rouleau in Quebec. These were mostly unaffected members of familial disease cohorts studied in different projects over the course of a decade^78,79^. The genomes of samples were sequenced with the Illumina platform at 30X average depth of coverage. The paired-end reads were mapped to GRCh38 (hg38) human genome assembly, and SNVs and indels were called using the Illumina DRAGEN pipeline (v3.5.3 and higher). We used only genotypes that pass all DRAGEN’s quality filters in the downstream analyses. Using our IBD analyses, we confirmed that these samples cluster together with QFC from CaG. To synthesize genotyping array data for the SNVs, indels, SVs, and HLA alleles imputation benchmarking, we subset WGS-based SNV and indel calls to genomic positions of variants on the Illumina GSA v2 array. We typed HLA alleles from WGS data using HLA*LA in the same way we did with CaG WGS data. We called SVs from WGS data the same way we did with CaG WGS data, the only difference being that we used Manta version 1.6.0 with default parameters instead of the DRAGEN implementation (see the corresponding Methods section). We used the “diploid” VCF output since the SVs were scored under a diploid model. Because we included only SVs with no more than 10% genotype missingness in the CaG reference panel, we applied the same filter on the called SV set for consistency.

#### Preparing SNVs, indels and SVs in WGS data for statistical phasing

We utilized WGS data from 2,173 individuals, including 1,756 QFC, 131 QMO, 163 QHA, and 123 additional participants of mostly European genetic ancestry to construct the genotype imputation reference panel. We replaced individual genotypes which did not pass variant calling quality filters from the Illumina DRAGEN pipeline (v3.9.5) with missing values (i.e. ‘./.’ genotypes). These missing genotypes were imputed during the subsequent statistical phasing. We left-aligned the variants using the vt algorithm^80^ and removed any resulting duplicated records. We recalculated the allele frequencies and removed monomorphic (alternate allele count is 0) variants. We added polymorphic high-quality SVs to the dataset (see the corresponding Methods sections on SVs). After these pre-processing steps, we had 70,047,094 variants (59,136,049 SNVs, 10,890,760 indels and 20,285 SVs) for subsequent statistical phasing.

#### Statistical phasing of SNVs, indels and SVs in WGS data

We used SHAPEIT5 (v5.1.1) statistical phasing software to build a CaG reference panel for imputing SNVs, indels and SVs^81^. To improve statistical phasing accuracy, we performed reference-based phasing with the harmonized high-quality reference haplotypes from the 1000 Genomes Project and Human Genome Diversity Project (1GP+HGDP) (N=4,904)^45^. First, we statistically phased genetic variants overlapping CaG and 1GP+HDGP with minor allele frequency (MAF) >0.1% in CaG to create a preliminary scaffold of CaG haplotypes of known common variants. Second, we statistically phased the remaining common CaG genetic variants (MAF>0.1%) not found in 1GP+HDGP using the preliminary scaffold from the first step to create a second scaffold of CaG haplotypes of all common variants. Lastly, we statistically phased the low-frequency CaG variants (MAF≤0.1%) using the scaffold from the second step to create a final set of CaG haplotypes. We performed statistical phasing in the pseudo-autosomal (PAR) regions in chromosome X in the same way as in all autosomal chromosomes. For the statistical phasing in non-PAR regions in chromosome X, we used the ‘--haploids’ option in SHAPEIT5 to provide male haploid sample identifiers.

#### Statistical phasing of HLA alleles in WGS data

We constructed a high-resolution HLA imputation reference panel combining the SNVs, indels, and HLA alleles from WGS data for 2,088 individuals using the HLA-TAPAS^27^ pipeline, which uses BEAGLE v4.1 to statistically phase together HLA alleles and single nucleotide polymorphisms (SNPs) restricting itself to the HLA region on chromosome 6. We used the HLA alleles called from WGS data which passed quality checks (43,150 alleles from 2,088 samples; see “*HLA genotyping*” section). For the SNV and indel data, we extracted the extended HLA region (chr6:25Mb–35Mb) from the WGS data after quality control (see “*Preparing SNVs, indels and SVs in WGS data for statistical phasing”)* for those individuals for whom HLA calling was available, and we excluded variants which were monomorphic (alternate AC<1) or with missingness greater than 0.1 in those individuals (228,651 SNVs and 59,903 indels).

#### Preparing genotyping array data for statistical phasing and imputation

We used the UCSC LiftOver tool^82^ to lift the data produced by each of the five genotyping arrays from the GRCh37 (hg19) to GRCh38 (hg38) human genome assembly. We removed lifted variant records that were not single nucleotide polymorphisms (SNPs), were palindromic, had none of the two alleles present in human genome reference, and were duplicated (keeping variant record with the lowest genotype missingness). We flipped the reported strand if necessary to align with the human genome reference. We set all heterozygous genotypes in the non-PAR region in chromosome X in male samples to missing values. Then, we merged the lifted data from the five genotyping arrays and removed genetic variants if they were missing in at least one of them. If the same sample was genotyped using multiple arrays, we kept only the version with the most genotyped positions. After merging, we removed SNPs that displayed statistically significant differences in alternate allele frequencies (AF) across genotyping arrays. We performed a likelihood ratio test (LRT) comparing two linear regression models G ∼ m + PC1 + … + PC4 + A2 + … + A5 and G ∼ m + PC1 + … + PC4, where G - individual genotype (0, 1, or 2 alternate allele copies), m - intercept corresponding to the average AF, PCi - principal component i, Aj - indicator variable for array j. Assuming random sampling and no batch effects, the array variables Aj should not impact the prediction of individual-specific AF. We computed the LRT P-value from a Chi-squared distribution with four degrees of freedom and set the statistical significance threshold of 0.05 divided by the number of tested SNPs. We obtained PCs using TRACE (v1.03)^83^ by projecting genotype array data into the PC space of the 1GP+HGDP reference panel. When performing LRT, we ignored all related individuals with ≤3rd degree of relatedness (PLINK’s --king-cutoff option set at 0.0442). We performed LRT for both sexes separately for variants in the non-PAR region in the X chromosome.

#### Statistical phasing of genotyping array data before imputation

We statistically phased the genotyping array data on 29,333 samples in CaG using the TOPMed r3 reference panel through TOPMed Imputation Server^73^, which uses Eagle2 (v2.4.1) for the reference-based statistical phasing^84^. Due to the maximal sample size restrictions in the TOPMed Imputation Server, we randomly allocated samples into two non-overlapping batches with N=14,666 (472,467 genotyped non-monomorphic variants) and N=14,667 (472,396 genotyped non-monomorphic variants) samples each for statistical phasing. The TOPMed Imputation Server’s quality checks removed 231 variants from each batch due to allele mismatch with TOPMed r3; 28,530 and 28,460 variants that were not present in TOPMed r3, respectively; 3 variants from each batch with genotype call rate <90%; 4 and 3 multi-allelic variants, respectively. The statistical phasing in non-PAR regions of chromosome X was done separately from the autosomal chromosomes and chromosome X PAR regions. We coded genotypes of male samples in chromosome X non-PAR regions as haploids, i.e. 0s and 1s instead of 0/0 and 1/1. After the statistical phasing in each batch, we merged all samples back together using the bcftools merge command^71^.

#### Genotype imputation of the pre-phased genotyping array data

We separately imputed genotype array data on 29,333 samples in CaG, which was pre-phased using the TOPMed r3 reference panel, into the CaG reference panel and TOPMed r3 reference panel. Then, we merged these imputation results using a meta-imputation approach. We used Minimac4 (v4.1.6)^85^ to locally impute the pre-phased array data into the CaG reference panel with default parameters and without explicitly chunking the chromosomes. The genotype imputation in non-PAR regions of chromosome X was done separately from the autosomal chromosomes and chromosome X PAR regions. We coded statistically phased genotypes of male samples in chromosome X non-PAR regions as haploids, i.e. 0s and 1s instead of 0|0 and 1|1. We utilized the TOPMed Imputation Server (v2.0.0-beta2), which also used Minimac4 (v4.1.6), to impute the pre-phased array data into the TOPMed r3 reference panel. Due to the server’s maximal sample size restriction, we performed imputation in two non-overlapping batches (see statistical phasing in Methods). We concatenated the imputed dosages from two batches and recalculated Rsq imputation qualities using the post-processing hds-util tool for Minimac4, which was developed by Michigan Imputation Server^85^. The chromosome X processing and male sample coding were identical to the local imputation into the CaG reference panel. We combined the imputed genotype dosages from the two reference panels using a meta-imputation approach implemented in MetaMinimac2 (v1.0.0)^86^.

### Genome-wide association study (GWAS) using imputed genotypes in CaG

#### Relatedness and population structure modelling

Our association models used genotyping array data to account for the relatedness between individuals and for population stratification. Using PLINK2 (v2.00a6LM)^87^, we selected only bi-allelic single nucleotide polymorphisms (SNPs) with minor allele frequency (MAF) >1%, Hardy-Weinberg test P-value <10^−15^, per-variant and per-sample missing genotype frequencies <10%. We further pruned SNPs based on the linkage-disequilibrium (LD) using the PLINK2 --indep-pairwise option with the window size of 1000 kbp, step size of 100 SNPs, and r^2^ LD threshold of 0.9 (i.e. two SNPs with r^2^>0.9 were considered in LD). We also removed SNPs in known long-range LD regions^88^ and low-complexity regions defined in UCSC Genome Browser (RepeatMasker and WM+SDust tracks)^89^. We performed principal component analyses (PCA) on pruned genotyping array data using the PLINK2 --pca option and extracted the top 10 principal components (PCs). We performed this variant pruning procedure and PCA on genotyping array data for all 29,333 individuals and 27,036 individuals of European-genetic ancestry separately.

#### Single variant association testing

For single variant genome-wide association testing between the imputed genotypes and continuous traits in CaG, we applied a linear regression model implemented in regenie (v3.6)^90^. We applied rank-inverse normal transformation (RINT) for all traits using the built-in regenie option during the estimation of polygenic effects on each trait (regenie Step 1) and the association testing (regenie Step 2). To estimate polygenic effects, we used pruned genotyping array data. We included sex, age, age^2^, genotyping array, and top 10 PCs as covariates to our regression models. Imputed variants with minor allele count (MAC) <5 were not considered. Subsequent filters on genotype imputation quality (INFO score reported by regenie) and minor allele frequency/count were applied and described in downstream analyses when required.

#### Genome-wide statistically significant loci, lead and index variants

We used a commonly accepted Bonferroni-corrected P-value threshold of 5×10^−8^ for the genome-wide statistical significance. We determined independent genome-wide significant loci, lead and index variants for each analyzed continuous trait separately. We used only genetic variants with the minor allele count (MAC) >20 and genotype imputation quality INFO score >0.3. First, we defined a window spanning 500 Kbp upstream and 500 Kbp downstream around each genome-wide significant association for a trait. Then, we merged all overlapping or immediately adjacent windows into a single independent genome-wide significant locus, which doesn’t overlap with any other locus for that trait. Within each independent genome-wide significant locus, we defined a single lead variant as the variant with the lowest P-value from the association test and one or more index variants as those which showed statistically significant associations using stepwise conditional analysis within each independent locus. We did stepwise conditional analyses with regenie (v3.6) using the same polygenic effects and covariates as in the primary genome-wide scan and defined the locus-wide statistical significance P-value threshold as 10^−5^.

### Replication of CaG variant-trait associations

We aimed to replicate the variant-trait pairs of interest in an independent dataset, the comprehensive cohort of the Canadian Longitudinal Study on Aging (CLSA) and the UK Biobank. To replicate associations with coding variants in the UK Biobank, we queried the Genebass server^51^. The CLSA is a longitudinal cohort of 26,622 females and males with genetic data and various phenotypes (questionnaires and lab measurements) aged 45 to 85 when recruited between 2010 and 2015 from across Canada^91,92^. To maximize sample size, we considered phenotypic information from the baseline visit. For sample quality control, we excluded participants identified as having batch or sex discordance between self-reported or chromosomal sex (or missing values for self-reported or chromosomal sex) or outlying heterozygosity as previously described^91^. In total 66 participants were excluded. We confirmed that the variants of interest for replication were in Hardy-Weinberg Equilibrium (P-value >1×10^−6^) in the CLSA dataset.

In the replication analysis, we included CLSA participants who passed quality control and were classified as having European genetic ancestry (N=24,655 samples) as previously described^91^. In brief, ancestry estimation was carried out by applying K-means clustering on principal components using Phase 3 of the 1000 Genomes Project samples as a reference. Given that a subset of CLSA participants were also recruited from the province of Quebec, we assessed kinship coefficients between the CLSA Quebec subset and CaG using KING to ensure independence of samples between the cohorts. We consequently removed 240 CLSA participants with a kinship coefficient with a CaG participant denoting a monozygotic twin or duplicate (> 0.354).

For the quantitative traits of interest, using R v4.3.1, we identified and then subsequently removed individuals with outlying value for the trait of interest, specifically those with values less than or greater than 5 standard deviations from the mean value for the trait. We carried out genome-wide association analyses using Regenie v3.2.1^90^ using the following covariates: genotyping batch, age, age squared, genetic sex, and the first 10 principal components of European ancestry. Rank inverse normal transformation was applied to the quantitative phenotypes. Samples that had missing information regarding phenotype information or covariates were removed. We consequently had up to 23,752 individuals of European genetic ancestry (of which up to 4,602 individuals were recruited from the province of Quebec). The alternative allele was tested as the effect allele as what was done in the CaG analyses. Variants with a nominal association P-value <0.05 in our CLSA replication analyses and exhibiting the same direction of effect as in the CaG association analyses were considered as successfully replicating.

### eQTL analyses

RNAseq data from whole blood was previously generated for 911 CaG participants^54^. We mapped reads onto the GRCh38 human transcriptome, derived from Ensembl v98 gene definitions, using kallisto v0.46.1 with the rf-stranded mode to get pseudoaligned reads to isoforms^93^. We obtained gene counts using the tx2gene function from the tximport R package^94^. The RNAseq libraries were processed with the edgeR and sva R packages^95,96^. The batch effect correction was assessed with ComBat, from the sva package, and genes having less than an average LCPM value of 0 were removed, resulting in a set of 13,641 genes. The 60 PEER factors were obtained from the normalized LCPM gene counts, according to the recommendations of the GTEx consortium^97^. After this step, two outliers’ samples were identified, and PEER was run a second time.

After quality-control steps, we retrieved 504 CaG participants with RNAseq and WGS data, from which 496 are QFC, 5 QHA and 3 QMO. Using PLINK v1.9, we removed variants with missing rate >1%, MAF <1% and a Hardy-Weinberg P<1×10^−6^. The low complexity regions were also removed, resulting in a total number of 11,164,382 variants. TensorQTL v1.0.8^98^ was run on a gene expression dataset that was used for the PEER correction and for which genes were subsequently inverse normal transformed using the cis-nominal mode. The linear regressions were corrected for age, age-squared, genetic sex, the 10 first PCs of the population structure, the WGS sequencing batch and 60 PEER factors. The gene expression data were further annotated with additional information, including the transcription start sites of the genes, based on the reference genome used to derive the gene counts (Ensembl rel. 98, GRCh38).

## DATA AVAILABILITY

The CaG genetic and phenotypic data is available from https://cartagene.qc.ca/en/researchers/access-request.html after scientific and ethical review. The CaG allele frequency information is publicly available from: https://cartagene-bravo.cerc-genomic-medicine.ca/. The GWAS and eQTL results are available from https://cerc-genomic-medicine.ca/pheweb/cartagene/ and https://eqtlbrowser-cag.mhi-omics.org/, respectively. Data are available from the Canadian Longitudinal Study on Aging (www.clsa-elcv.ca) for researchers who meet the criteria for access to de-identified CLSA data.

## CODE AVAILABILITY

The source code used to process the data and generate data for tables and figures is available through figshare at https://doi.org/10.6084/m9.figshare.28891670.

## ACKNOWLEDGMENTS

This research has been conducted using Data and Biosamples from CARTaGENE (https://cartagene.qc.ca/en). This work has been made possible through financial support from Genome Québec (Ministère de l’Économie, de l’Innovation et de l’Énergie), the Canadian Partnership Against Cancer and Health Canada. Grants from Genome Quebec, Genome Canada and CHU Ste-Justine supported the GenoRef-Q whole-genome sequencing project. This research was enabled in part by support provided by Calcul Québec (https://www.calculquebec.ca/) and the Digital Research Alliance of Canada (alliancecan.ca). Work in the Lettre lab is funded by the Canadian Institutes of Health Research (Projects #426541 and #486808), the Canada Research Chair program, and the Montreal Heart Institute Foundation (G.L.); Fondation Courtois and FRQS-Junior 2 (M.T.); Work in the Gravel lab was funded by the Canada Research Chair program, CIHR project grant 437576 and NSERC discovery grant; S.A.G.T. was funded by a Junior 2 award from the Fonds de Recherche du Québec - Santé (FRQS; https://frq.gouv.qc.ca). Work in the Gagliano Taliun lab is funded by the Alzheimer Society Research Program by the Alzheimer Society of Canada and co-funding by the Canadian Institutes of Health Research – Institute of Aging (Funding Reference Number 189919), as well as by additional support by the Canadian Institutes of Health Research (PJT 183817, PJT 191687 and AD6 192920); FRQS-Junior 1 (C.B.); FRQS Senior (A.-M.L.); National Institutes of Health grant numbers R35-CA197449, R01-HL163560, U01HG012064 (H.Z.). J.H. was a FRQS Junior 2 fellow, work was funded by Montreal Heart Institute Foundation, NSERC Discovery Grant (RGPIN-2022-04262) (J.H.). This research was made possible using the data/biospecimens collected by the Canadian Longitudinal Study on Aging (CLSA). Funding for the Canadian Longitudinal Study on Aging (CLSA) is provided by the Government of Canada through the Canadian Institutes of Health Research (CIHR) under grant reference: LSA 94473 and the Canada Foundation for Innovation, as well as the following provinces, Newfoundland, Nova Scotia, Quebec, Ontario, Manitoba, Alberta, and British Columbia. This research has been conducted using the CLSA datasets: baseline comprehensive dataset – version 7.0 and Genome-wide genetic data – version 3.0 under Application Number 23ME002. The CLSA is led by Drs. Parminder Raina, Christina Wolfson and Susan Kirkland. The opinions expressed in this manuscript are the author’s own and do not reflect the views of the Canadian Longitudinal Study on Aging.

## AUTHOR CONTRIBUTIONS

Conceptualization of the project: D.T., G.L., S.Gravel; Funding acquisition: G.L., S.Gravel; Project management: A.M., C.L., D.T., G.L., S.Gravel; Oversight and leadership responsibility: A.-M.L., C.B., D.T., G.L., J.H., M.T., S.A.G.T., S.Gravel, S.Girard; Provided additional resources: D.S., G.R.; Developed software and websites: H.X., J.-C.G., V.C., J.H., D.T.; Data curation: J.-C.G., J.P., S. Gravel, D.T., V.C., G.L., G.M., J.-F.T., K.S.L., L.A.-T., P.M., R.L., T.d.M.; Method development and creation of models: J.-C.G., J.P., D.T., S. Gravel, G.L., K.S.L., L.A.-T., P.M., R.L., C.B., G.F., G.G., J.B., M.L., M.T., S.A.G.T., A.D.-P., A.D.M.; Data analysis: J.-C.G., J.P., D.T., S. Gravel, G.L., P.M., R.L., C.B., G.F., G.G., J.B., M.L., M.T., A.D.M., V.C., J.-F.T., T.d.M., A.M.-G., H.Z.; Writing, figure and table production, and revision: all authors.

