## Supplementary Notes and Figures for "A multi-ancestry genetic reference for the Quebec population"

**1. Glossary of population labels**

There is currently no consensus on the use of population descriptors in biomedical research in Quebec. In particular, some descriptors have been used interchangeably to refer to groups based on language, culture, or ancestry. Here we define terms as we will use them in this work.

- **Quebec Residents (QR)**: A person living in Quebec, Canada.
- **French-Canadian (FC)**: A term that is used variably in the literature to refer to linguistics (Canadians who speak French), ethnicity (an ethnic group with ties to French ancestry), and ancestry (individuals who trace their ancestry to New France settlers). Here we will use FC as an adjective to refer to ancestry from the French settlers.
- **Quebec residents of French-Canadian ancestry (QFC).** Individuals in Quebec with appreciable FC ancestry. Since we do not have direct access to genealogical ancestry for all individuals, we used the existing derived variable ETHNICITY_FR from the CARTaGENE cohort, which considers as likely QFC individuals who are born in Quebec, have two parents and four grandparents born in Canada, have first language French, and self-identified as white. In other words, inferred QFC status depends on both cultural and ancestral criteria, with the understanding that this will exclude QFC individuals who do not have French as a First language. It also includes individuals of Acadian ancestry.
- **Quebec Residents with Morocco-born grandparents** **(QMO)**: Quebec residents whose four grandparents were born in Morocco.
- **Quebec residents with Haiti-born grandparents (QHA)**: Quebec residents whose four grandparents were born in Haiti.
- **AFR**: Refers to a genetically-inferred ancestry component inferred by the Admixture software, which we interpret as reflecting ancestors living in Africa in 1450.
- **AMR**: Refers to a genetically-inferred ancestry component inferred by the Admixture software, which we interpret as reflecting ancestors living in the Americas in 1450.
- **EUR**: Refers to a genetically-inferred ancestry component inferred by the Admixture software, which we interpret as reflecting ancestors living in Europe in 1450.
- **NFE** (Non-Finnish European): a classification based on genetic similarity used by gnomAD to report allele frequencies across European-ancestry populations (https://gnomad.broadinstitute.org/news/2023-11-genetic-ancestry/).
- **Indigenous**: An ethnic label referring to the Inuit, First Nations, and Métis nations.
- **Québécois**: In English, as a noun, an ethnic label applying to the population of Quebec, often implying francophone culture or French ancestry. In French, it typically refers to all Quebec residents, as either a geographic or ethnic/nationhood classifier. While a commonly preferred ethnonym, we avoid it due to the mismatch between English and French definitions.
- **European genetic ancestry** (for genome-wide association studies); We selected CaG participants of inferred European genetic ancestry using the following steps: (1) We projected CaG samples to the reference PCA space constructed using unrelated individuals in the 1000 Genomes Project (1000G). (2) For each CaG individual, we computed the Mahalanobis distance to each of the 5 superpopulations in the 1000 Genomes Projec PCA space (Admixed American, African, East Asian, South Asian, European). We used the first 6 principal components and Mahalanobis.py (https://github.com/CERC-Genomic-Medicine/scripts/blob/master/Mahalanobis.py) to compute the Mahalanobis distance. (3) We assigned a CaG individual to a superpopulation if the corresponding Mahalanobis distance's P-value was ≥0.001 and all other P-values were <0.001. (4) We removed 879 CaG individuals for whom the inferred genetic ancestry did not match the ETHNICITY variable value derived from the CaG questionnaire data. (5) We confirmed that 25,017 CaG individuals with assigned European genetic ancestry: cluster together with European ancestry individuals in 1000 Genomes Project and have a vast proportion of European ancestry autosomal chromosomes.

**2. Identity-by-descent (IBD) cluster analysis**

We first detected pairwise identity-by-descent (IBD) segments among all CaG participants with genome-wide genotyping data using the software *hap-ibd* (<https://doi.org/10.1016/j.ajhg.2020.02.010>). Genotypes were quality-controlled and phased with *Beagle* 5.5 prior to IBD detection. We applied the default *hap-ibd* parameters as recommended in the documentation, which include a minimum seed segment length of 2.0 cM (min-seed=2.0), a maximum gap of 1000 base pairs between IBS segments for extension (max-gap=1000), and a minimum extension segment length of 1.0 cM (min-extend=1.0). Only output IBD segments of at least 2.0 cM (min-output=2.0) and with at least 100 markers (min-markers=100) were retained. Markers with a minor allele count below 2 were excluded (min-mac=2). Using these parameters, we computed the average total length of the IBD segments shared between all CaG participants with genome-wide genotyping data available (between and within IBD clusters, **Supplementary Fig. 2**). The average IBD sharing between clusters is 4.456 cM, although the average total length ranges from 2.19 to 32.49 cM in pairs of clusters. Clusters 1, 6, 7, 10, 17, 21, 30, 31, 34 and 35 show high within-cluster IBD and have Canada as the most frequent country of birth. In fact, more than 99.24% of the samples in these clusters were born in Canada, with 58 individuals with unknown country of birth out of the 15,907 individuals. Average IBD sharing within cluster ranged from 2.9 cM for pairs of samples in cluster 42 to 32.5cM in cluster 1. The mean length of shared IBD segments in cluster 1 is comparable to what would be expected from a single IBD segment inherited from a common ancestor shared by third cousins. Notably, 21.5% of individuals in cluster 1 were recruited in the Saguenay region, reflecting the well-known founder effect in this region of Quebec. Cluster 16 comes second with an average of 25.7cM. In this cluster, a high proportion of individuals have self-declared ethnicity including the term “Jewish”. This finding suggests that IBD clustering enables identifying this other founder population group in CaG. Cluster 30 comes third with an average pairwise IBD sharing of 22.4cM. Province of birth as well as self-declared ethnicity provided insight into the origin of individuals forming this cluster that we suspect are Acadians: most of the participants in this cluster are born in Quebec (85.10%), but 98 individuals were born in New-Brunswick, six in Nova-Scotia and three in Prince-Edward Island, representing 42.13% of all CaG participants born in these three Atlantic provinces. This hypothesis is coherent with the known founder effect for this population in the Atlantic provinces and in Gaspesia.

*A) Comparison with UMAP+HDBSCAN clustering*

We compared the 21 IBD clusters to the 14 HDBSCAN clusters (**Fig. 1B** and **Supplementary Fig. 3**). The group of clusters representing people born in Canada (IBD cluster 1, 6, 7, 10, 17, 21, 30, 31, 34 and 35) are almost all found in the HDBSCAN cluster 14, with the exception of cluster 1 and cluster 30, which respectively have 54% in HDBSCAN cluster 13 and 73% in HDBSCAN cluster 12. These two clusters are also the ones identified as “Saguenay'' and “Acadian” in the IBD analysis (see above). As for individuals in IBD clusters 40, 42, 43 and 55, they all cluster together in HDBSCAN cluster 1 corresponding to the Asian descent cluster. IBD clustering enables us to differentiate sub-population in Asia even with a small sample size. For example, 19 out of 23 individuals in IBD cluster 55 are born in the Philippines.

**3. Building a reference haplotype panel**

A set of high-quality reference haplotypes genetically similar to a study population, often referred to as a haplotype reference panel, is required for many genetic analyses, including genotype imputation, genetic ancestry inference, and estimation of linkage disequilibrium (LD) between variants in genome-wide association studies (GWAS). Using the high-depth WGS data from 2,173 CaG participants, we constructed a haplotype reference panel which included ~80M genetic variants. We assessed the accuracy of the statistical phasing (i.e., haplotype inference from genotypes) in the CaG reference panel by adding children from 60 independent family trios to the panel and re-phasing it, one at a time. The trio children of reported East Asia (EAS) and Central South Asia (CSA) genetic ancestries showed the highest average haplotype switch error rates (SER=2.721% [SE=0.010%] and SER=1.723% [SE=0.067%], respectively) compared to others, which we explain by the minimal number of individuals with related ancestries in the CaG panel. The SER values decreased as the number of individuals in the CaG panel who were genetically similar to the trio's children reported genetic ancestries grew. The lowest average SER values were in children of French-Canadian (QFC) genetic ancestry (SER=0.152% [SE=0.005%]), the largest ancestry group in the CaG panel, followed by the children of European (EUR, SER=0.518% [SE=0.019%]), African (AFR, SER=0.753% [SE=0.005%]), and Admixed American (AMR, SER=1.013% [SE=0.094%]) genetic ancestries (**Supplementary Fig. 12** and **Supplementary** **Table 10**). We then applied a reference-based statistical phasing, implemented in SHAPEIT5, that leveraged information from the external 1000 Genomes Project and HGDP reference panel, and re-evaluated the accuracy of the inferred haplotypes. The average SER values improved in all trio children (**Supplementary Fig. 13** and **Supplementary** **Table 11**). The largest absolute improvement was in the trio children of EAS genetic ancestry (from average SER=2.721% [SE=0.010] to 1.187% [SE=0.013%]; average paired difference SER=1.535% [SE=0.010%]), while the smallest absolute improvement was in the trio children of QFC genetic ancestry (from average SER=0.152% [SE=0.005%] to 0.147% [SE=0.005%]; average paired difference SER=0.005% [SE=0.001%]). We observed the improved average SER across all variants with at least two alleles in the CaG reference panel in all trio children except those of QFC genetic ancestry (**Supplementary Fig. 14** and **Supplementary** **Table 12**). In the trio children of QFC genetic ancestry, although the prior phase information from the external reference panel helped improve average SER values across most of the allele count spectrum, in many cases these improvements did not reach statistical significance (P-value <0.005 after adjusting for comparisons in 10 independent allele count bins).

**4. Evaluation of SV imputation quality**

In our independent benchmark dataset (N=141), using WGS data, we identified 5,657 SVs (5,305 DEL and 352 DUP) after quality filters (7,182 SVs before quality filters). Then, we aimed to compare the concordance between WGS-based SVs and SVs obtained through imputation from the CaG reference panel. For the imputed SV set, we looked at three subsets based on the imputation R^2^: SVs with R2 ≥ 0.75 (6,604 SVs), R2 ≥ 0.30 (7,625 SV), and 0.75 > R2 ≥ 0.30 (1,561 SVs). Since PLINK (the --pgen-diff function) needs the positions and identifiers (IDs) of two variants to be identical for the genotype concordance to be calculated, we first had to intersect the called SV set and the imputed SV set to find the pairs of SVs that would be considered the same. To do so, we used bedtools version 2.31.0 to intersect the sets of SVs with a 50% reciprocal overlap. We created new IDs unique for a pair and changed the position of one SV of the pair if needed in Python and calculated the concordance of genotypes with PLINK for each of the subsets. Deletions and duplications were analyzed separately. For the subset of SVs with R^2^ ≥ 0.75, the genotype concordance reached 97.82% (98.50% in autosomal chromosomes only) and 98.81% for 3,728 deletions and 169 duplications compared, respectively. For the subset of SVs with R^2^ ≥ 0.30, the concordance reached 96.48% (98.41% in autosomal chromosomes only) and 98.83% for 3,969 deletions and 175 duplications compared, respectively. Lastly, for the subset of SVs with 0.75 > R^2^ ≥ 0.30, the concordance was 75.37% (96.33% in autosomal chromosomes only) for 241 deletions compared.

**5. GWAS results for structural variants (SVs)**

In the cross-ancestry GWAS for quantitative traits, we found 42 significant associations with SVs (0.05/15,000 SV tested = 3.3x10^-6^), including 21 genome-wide significant SVs (*P*<5x10^-8^, **Supplementary Table 15**). The 15+kb French-Canadian deletion of the *LDLR* gene (minor allele frequency [MAF]=0.0014) is significantly associated with an increase in LDL-C and total cholesterol with respective P-values of 1.6x10^-9^ and 7.5x10^-9^. We also found an 8.3kb deletion upstream of *NEGR1* associated with decreased body mass index. It has also been reported that a 8-kb deletion upstream of *NEGR1* encompassing a conserved transcription factor binding site for *NKX6.1* is protective against obesity^1^. Very similar SV associations have been reported before for nine of our significant SV associations:

Two common deletions (located 200-kb and 350-kb upstream of *SLC2A9*) have been previously associated with serum uric acid concentration: chr4:10,221,780-10,233,632, an 11.8-kb deletion with a decrease, and chr4:10,391,518-10,399,032, a 7.5-kb deletion associated with an increase in serum uric acid concentration^2^. In CaG, we found a 23-kb deletion (chr4:10,209,636-10,232,944) associated with decreased serum uric acid overlapping the 11.8-kb deletion, as well as a 9.8-kb and 9.7-kb deletions (chr4:10,390,708-10,400,558 and chr4:10,390,810-10,400,543) associated with increased serum uric acid completely overlapping the previously described 7.5-kb deletion.

SVs have been associated with hematological traits in the NHLBI TOPMed program^3^, and some of the reported associations were also found in our GWAS. We found a 589-bp deletion (chr6:41,897,029-41,897,618) associated with decreased MCH and MCV while a 538-bp deletion associated with decreased MCH and MCV (chr6:41,897,089-41,897,626) was reported by the TOPMed program. We identified a 3.46-kb deletion (chr6:41,985,492-41,988,955) and a 3.29-kb deletion (chr6:41,985,600-41,988,895) in the intron of *CCDN3* associated with increased MCV and MCH, while a 3.31-kb deletion (chr6:41,985,574-41,988,887) in the intron of *CCND3* associated with MCV was reported in the TOPMed program. We found a 54-bp deletion (chr6:31,132,407-31,132,461) in an intron of *PSOR1C1* associated with increased lymphocyte counts while a 57-bp deletion (chr6:31,132,409-31,132,465) was reported in the TOPMed program. And finally, a 3.72-kb deletion in the intron of *ATXN2* (chr12:111,538,485-111,542,205) associated with decreased platelets while a deletion associated with the same breakpoints was reported in TOPMed.

We performed conditional analysis for the 42 significant SVs conditioning on the most significant SNP in a 1-Mb window (SNPs with MAC > 5). 6 SVs were already the most or one of the most significant variants in the window. However, these variants have a MAF <0.4% and needs to be replicated in independent studies (**Supplementary Table 15**). Our conditional analyses indicated that for all SVs, SNPs were either equally good or better predictors of the phenotypic variation.

**6.** **Evaluation of HLA allele imputation quality**

The CaG HLA panel was used to impute HLA alleles into an independent validation cohort of synthesized genotype array data from 141 QFC individuals. We imputed HLA alleles into the simulated genotype array data and compared concordance to those inferred in the WGS data. Alleles imputed by the CaG HLA panel were in good concordance with HLA alleles typed from WGS data, even up to high (G-group/three-field) resolution (97.0% 96.3%, and 96.2% of alleles were correctly-imputed at one-field, two-field, and G-group allele resolution, respectively). We tested multiple thresholds for filtering on imputation quality (R^2^) and set a quality threshold of R^2^>0.6 for imputed alleles, removing those low-confidence imputed alleles which fell below this threshold. After filtering, concordance of imputed alleles with WGS was improved at each resolution, to 99.1%, 98.7%, and 98.6% of alleles concordant-per-sample, respectively.

**7.** **GWAS results with imputed HLA alleles**

Using imputed HLA alleles, we performed association testing with 42 continuous health-related traits in two subsets: European genetic ancestry individuals (N=25,210) and all ancestries (N= 27,239). 47 statistically significant associations were detected (P-value <3.7x10^-4^) in the multi-ancestry analysis across 20 phenotypes (**Supplementary Table 16**). 38 of these associations were also present in the European genetic ancestry analysis, while an additional eight hits were only significant in Europeans. Of 137 unique two-field HLA alleles imputed in CaG, 32 were statistically significantly associated with at least one trait in the entire cohort and in the European-ancestry subset. Eight of the 10 most significant hits were with hematological traits. White blood cell (WBC) count and TSH each were significantly associated with six alleles (two shared alleles were associated with both phenotypes) (**Supplementary Fig. 18**). The two strongest associations were with HLA-B*08:01, with monocyte count (adjusted P-value=4.85E-10, effect size=-0.021, SE=0.0034) and WBC count (p-value=8.43E-08, effect size=-0.196, SE=0.037) (**Supplementary Fig. 18**). These findings were also consistent in CaG participants with genetic European ancestry (P-value=5.51E-10, effect size=-0.022, SE=0.0035, and P-value=1.47E-07, effect size=-0.198, SE=0.037, respectively). B*08:01 was also significantly associated with lymphocyte count (P-value=2.4E-05, effect size=-0.056, SE=0.013) and with thyroxine (T4) levels (P-value=3.55E-05 effect size=0.146, SE=0.035), both in the cross- and European-ancestry groups. HLA-B*08:01 has previously been associated with several HLA-related traits, particularly with elevated plasma beta-2-microglobulin levels^4^, which have also been shown to associate with elevated thyroid traits, including thyroxine^5^, as found in thyroid disease such as Graves’ disease. Other HLA phenome-wide association studies conducted in population biobanks, FinnGen^6^ and UK Biobank^7^, reported associations between HLA-B*08:01 and several phenotypes including thyroid disease. In CaG, precise phenotypic health measurement data enabled us to pinpoint the specific effect direction (i.e. elevated thyroxine levels) and pleiotropic effects with multiple blood measurements. HLA-B*08:01 is known to be located on a European-ancestry haplotype, which was confirmed by our finding that its allele frequency is higher in QFC (AF=0.09) compared to QHA (AF=0.02) and QMO (AF=0.05). It was also detected at higher frequency (AF=0.09) in the CaG cohort compared to a global multi-ancestry reference panel (AF=0.068)^8^.

**Supplementary Table 1. Brief description of the CARTaGENE (CaG) cohort.** QFC, Quebec residents of French-Canadian ancestry. For more details about the design and demographic characteristics of CaG, see Awadalla et al.^9^

|  | **Whole CaG cohort** | **Genotyped**  **CaG subset** | **Whole genome sequenced**  **CaG subset** |
| --- | --- | --- | --- |
| **Sample size (N)** | 43,032 | 29,337 | 2,184 |
| **Women (%)** | 23,426 (54%) | 15,691 (53%) | 1,188 (54%) |
| **Mean age at baseline (SD)** | 54 (±8) | 54 (±8) | 54 (±8) |
| **QFC (%)** | 26,305 (61%) | 18,871 (64%) | 1,756 (80%) |
| **Participants with 4 Morocco-born grandparents (%)** | 302 (0.70%) | 201 (0.68%) | 132 (6.0%) |
| **Participants with 4 Haiti-born grandparents (%)** | 367 (0.85%) | 270 (0.92%) | 163 (7.5%) |

**Supplementary Table 4. A non-exhaustive list of ClinVar-defined pathogenic variants that are at least 4-times more frequent in the CARTaGENE (CaG) cohort than in matched populations from the gnomAD database.** For the variants identified in CaG French-Canadian (QFC), Haitian (QHA) and Moroccan (QMO) participants, we used allele frequencies from gnomAD non-Finnish European, African and Middle East populations, respectively.

| **Variant_Gene** | **CaG_MAF** | **Enrichment ratio (CaG/gnomAD)** | **Disease** |
| --- | --- | --- | --- |
| ***QFC*** |  |  |  |
| rs74315323_*HJV* | 0.0026 | 5 | Hemochromatosis type 2A. |
| rs119466000_*LRPPRC* | 0.0020 | 136 | Congenital lactic acidosis, Saguenay-Lac-Saint-Jean type. |
| rs119478059_*MRPS22* | 0.0014 | 5 | Hypotonia with lactic acidemia and hyperammonemia. |
| rs199815268_*SGO1* | 0.0071 | 28 | Chronic atrial and intestinal dysrhythmia. |
| rs606231134_*SYNE1* | 0.0028 | 39 | Autosomal recessive ataxia, Beauce type. |
| rs119103243_*SYNE1* | 0.0017 | 116 | Autosomal recessive cerebellar ataxia, Beauce type. |
| rs111033307_*SLC26A4* | 0.0023 | 14 | Rare genetic deafness (Pendred syndrome). |
| rs77188391_*CFTR* | 0.0020 | 68 | Cystic fibrosis. |
| rs118204060_*LPL* | 0.0014 | 48 | Hyperlipoproteinemia, type I. |
| rs755595256_*PLPBP* | 0.0020 | 136 | Epilepsy, early-onset, vitamin B6-dependent. |
| rs121908012_*UROS* | 0.0028 | 8 | Cutaneous porphyria. |
| rs61754361_*TYR* | 0.0017 | 39 | Tyrosinase-negative oculocutaneous albinism. |
| rs387906260_*CYP27B1* | 0.0031 | 43 | Vitamin D-dependent rickets, type 1A. |
| rs281865117_*SACS* | 0.0069 | 233 | Charlevoix-Saguenay spastic ataxia |
| rs28928906_*MPI* | 0.0020 | 136 | MPI-congenital disorder of glycosylation |
| rs80338901_*FAH* | 0.0054 | 11 | Tyrosinemia type I. |
| rs113994205_*CTNS* | 0.0020 | 10 | Nephropathic cystinosis. |
| rs121434372_*GCDH* | 0.0028 | 11 | Glutaric aciduria, type 1. |
| rs606231167_*PNPLA6* | 0.0014 | 5 | Hereditary spastic paraplegia. |
| ***QHA*** |  |  |  |
| rs116807569_*AGPAT2* | 0.0061 | 4 | Congenital generalized lipodystrophy type 1. Identified in families of African descent (OMIM 603100). |
| rs140291094_*STAC3* | 0.0061 | 6 | Bailey-Bloch congenital myopathy. Identified in patient of Afro-Carribean descent (OMIM 615521). |
| ***QMO*** |  |  |  |
| rs397704718_*FAM161A* | 0.0076 | Absent from gnomAD | Retinitis pigmentosa 28. Identified in Moroccan Jewish families (OMIM 613596). |
| rs397515377_*TTPA* | 0.0076 | Absent from gnomAD | Ataxia, Friedreich-like, with isolated vitamin E deficiency. Families from North Africa and Italy (OMIM 600415). |
| rs267606777_*DSG4* | 0.0076 | Absent from gnomAD | Hypotrichosis. Identified in Moroccan and Iraqi Jewish families (OMIM 607892). |

**Supplementary Table 8. Allele frequency of pathogenic and likely pathogenic variants in *SPG7* across CARTaGENE (CaG) subgroups.** Each row is color-coded based on classification of pathogenicity in ClinVar: pathogenic/likely pathogenic variants are in pale green, variants of unknown significance or with conflicting interpretations of pathogenicity are in orange, and variants not included in gnomAD and ClinVar are in white. NA, not available.

|  | | | | **CaG WGS** | **Allele frequency (AF) in CaG WGS** | | | **gnomAD AF** |
| --- | --- | --- | --- | --- | --- | --- | --- | --- |
| **Variant** | **rsID** | **GRCh38** | **Consequence** | **Alternate Allele Count** | **QFC** | **QHA** | **QMO** |  |
| c.1529C>T (p.A510V) | rs61755320 | 16:89546737 C>T | Missense | 21 | 0.0051 (1/195) | 0 | 0.0038 (1/262) | 0.005795 |
| c.988-1G>A | rs748309520 | 16:89531903 G>A | Splicing | 3 | 0.0009 (1/1171) | 0 | 0 | 0.000009913 |
| c.1045G>A (p.G349S) | rs141659620 | 16:89531961 G>A | Missense | 4 | 0.0011 (1/878) | 0 | 0 | 0.001589 |
| c.1715C>T (p.A572V) | rs72547551 | 16:89550545 C>T | Missense | 2 | 0.0006 (1/1756) | 0 | 0 | 0.00006815 |
| c.1940C>T (p.A647V) | rs776380988 | 16:89553797 C>T | Missense | 2 | 0.0006 (1/1756) | 0 | 0 | 0.000001240 |
| c.25C>T (pR9C) | rs1368314619 | 16:89508442 C>T | Missense | 2 | 0.0006 (1/1756) | 0 | 0 | 0.00002530 |
| c.2293G>A (p.D765N) | rs374941242 | 16:89556998 G>A | Missense | 3 | 0.0009 (1/1171) | 0 | 0 | 0.00002231 |
| c.376+1G>T | rs746053679 | 16:89513038 G>T | Splicing | 2 | 0.0006 (1/1756) | 0 | 0 | 0.00001122 |
| c.2103+2T>C | NA | 16:89553962 T>C | Splicing | 1 | 0 | 0.0031 (1/326) | 0 | NA |
| **Cumulative AF for this set of nine pathogenic/likely pathogenic variants** | | | | | 0.0103 (1/98) | 0.0031 (1/326) | 0.0038 (1/262) |  |

**Supplementary Table 9.** **Number of carriers for variants in the *SPG7* gene**. We estimated carriers directly from WGS data, and using Ancestral recombination graph (ARG)-based imputation for eight variants in the *SPG7* gene. The number of samples with WGS data is 2173 and the number of imputed samples is 29,337. We used ARG-based estimation of the time to the most recent common ancestor (TMRCA)^10^.

|  | **Number of QFC carriers** | |  |
| --- | --- | --- | --- |
| ***SPG7***  **variant** | **WGS** | **ARG-imputation** | **TMRCA (generations)** |
| c.1045G>A (p.G349S) | 4 | 45 | 12.86 |
| c.1529C>T (p.A510V) | 20 | 244 | 31 |
| c.1715C>T (p.A572V) | 2 | 7 | 5.48 |
| c.1940C>T (p.A647V) | 2 | 4 | 4.41 |
| c.25C>T (pR9C) | 2 | 12 | 8.76 |
| c.2293G>A (p.D765N) | 3 | 46 | 14.38 |
| c.376+1G>T | 2 | 6 | 7.7 |
| c.988-1G>A | 3 | 44 | 8.01 |

**Supplementary Figure 1.** **Data reduction and visualization for the CaG participants with genome-wide genotyping data available.** PC4 vs PC3 (left) and UMAP (right). Clusters are determined by UMAP and HDBSCAN as described in **Methods**. Word clouds are provided for country of birth, province of birth (for those born in Canada), collection region, and selected ethnicity. Word clouds for cluster 6 have been excluded because of its small size (N<30).

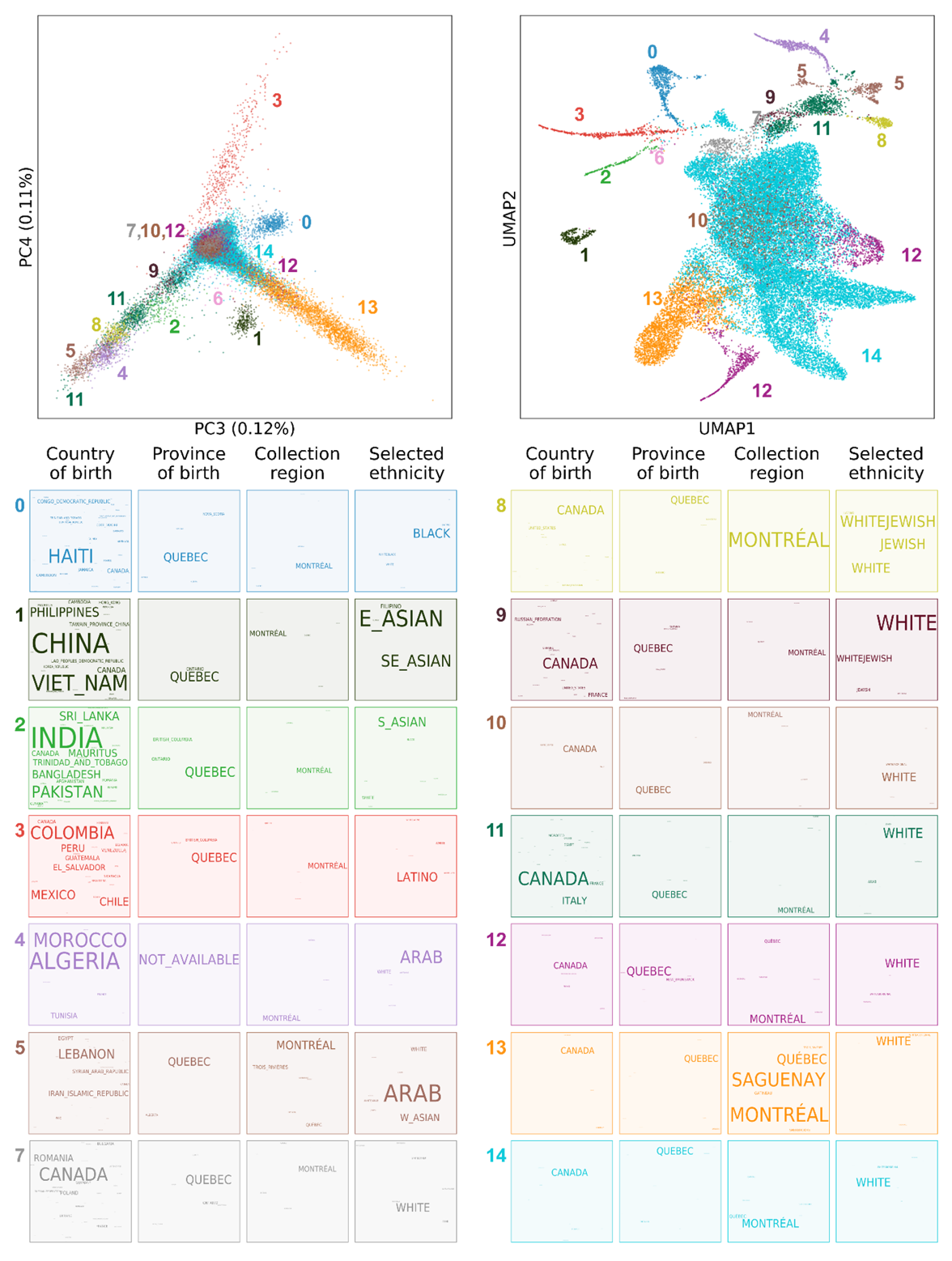

**Supplementary Figure 2. Average identity-by-descent (IBD) sharing between and within clusters in 29,209 CARTaGENE (CaG) participants.** Left. Heatmap of the average total length of IBD sharing segments in cM between pairs of individuals from the IBD clusters. Right. Boxplot of the average total length of IBD segment sharing between individuals within each cluster. See text above in the **Supplementary Notes** for details.

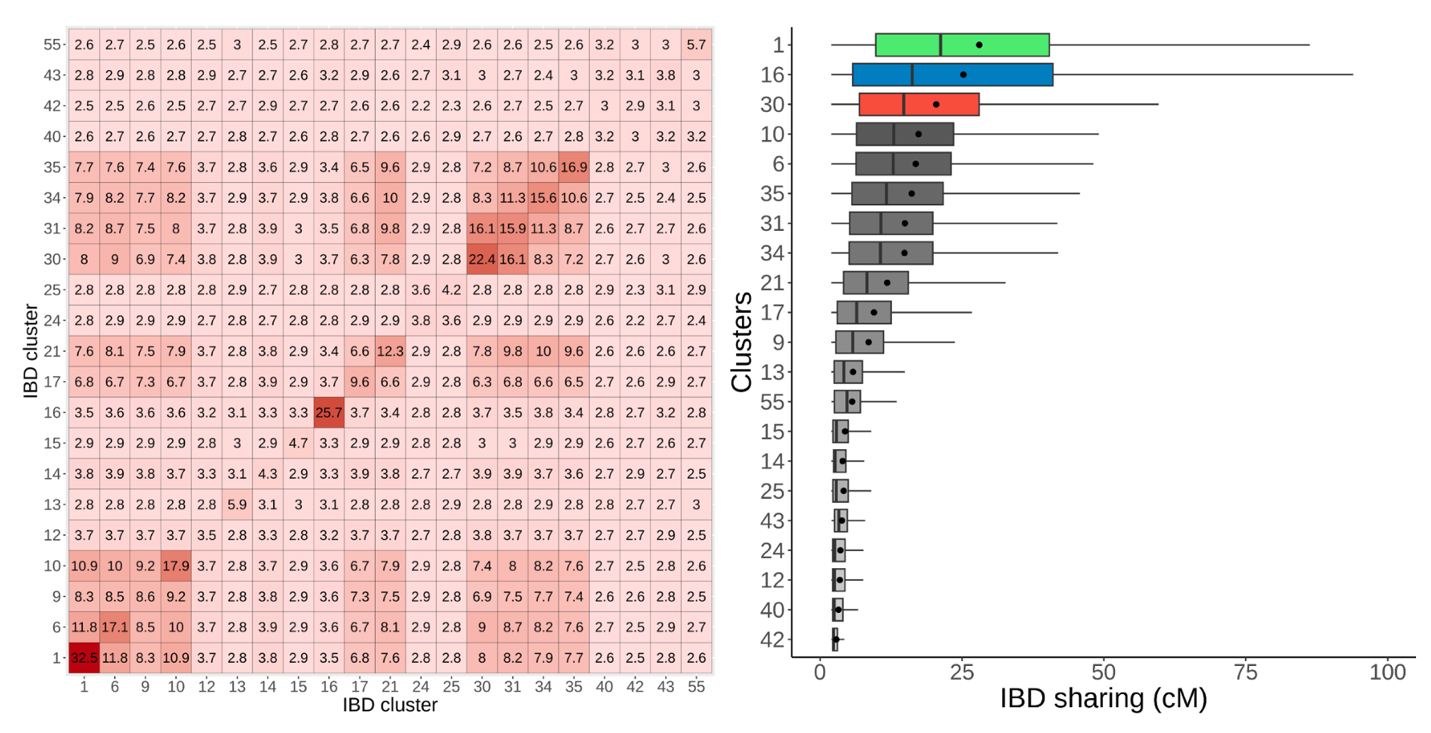

**Supplementary Figure 3. Comparison of IBD clustering and HDBSCAN clustering.** Proportion of samples in each IBD cluster that are found in the HDBSCAN clusters (rounded to 2 digits)

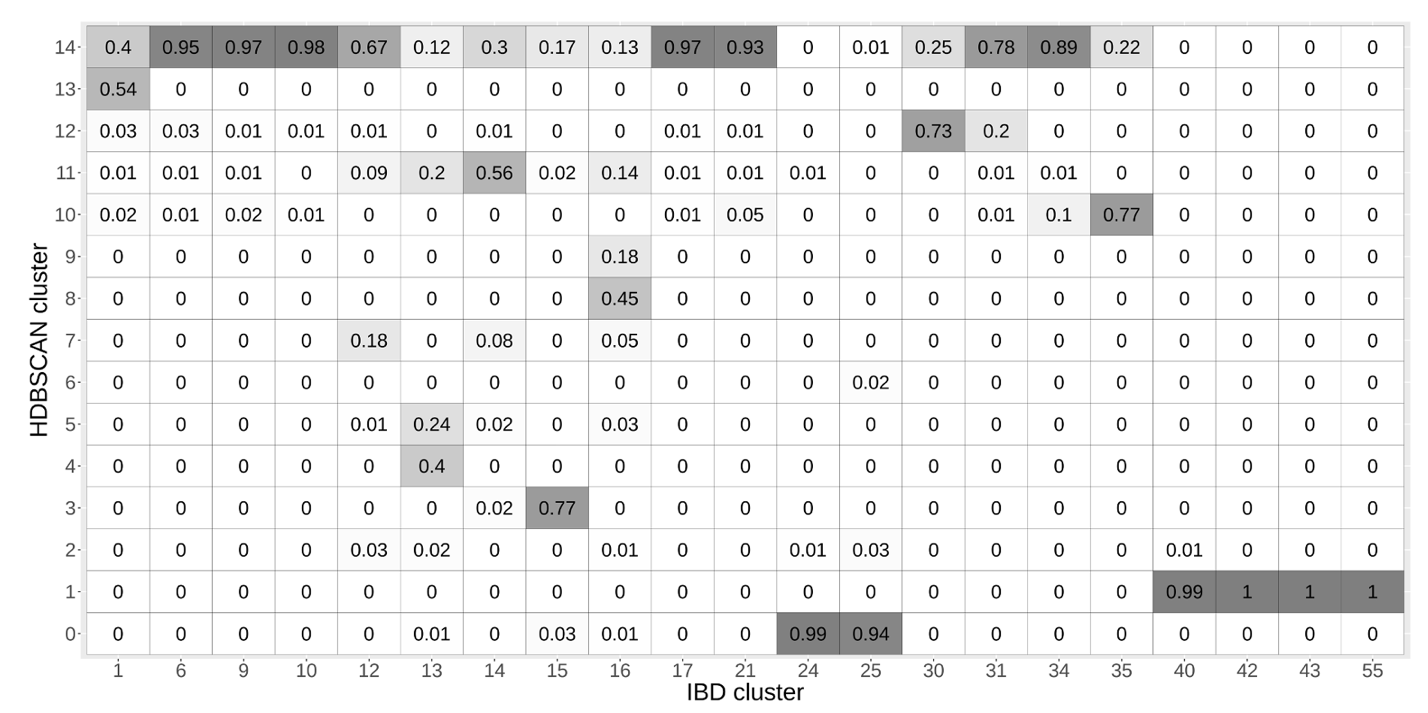

**Supplementary Figure 4. Admixture analysis of CaG participants with four grandparents born in Morocco.** (**A**) Estimation of admixture proportions among CaG participants whose four grandparents were born in Morocco (QMO) for K=3 to K=7. We included as reference CaG participants with four grandparents born within the same country in North Africa, Sub-Saharan Africa, and Europe. (**B**) Immigration year by QMO admixture cluster portraying only individuals born in Morocco (N=194). Cluster 1 (green, left), predominantly “Arab” as self-declared ethnicity (N=163, mean=1998, standard deviation [sd]=9). Cluster 2 (orange, right), predominantly “Jewish” self-declared ethnicity, not counting NAs (n=31, mean=1977, sd=11).

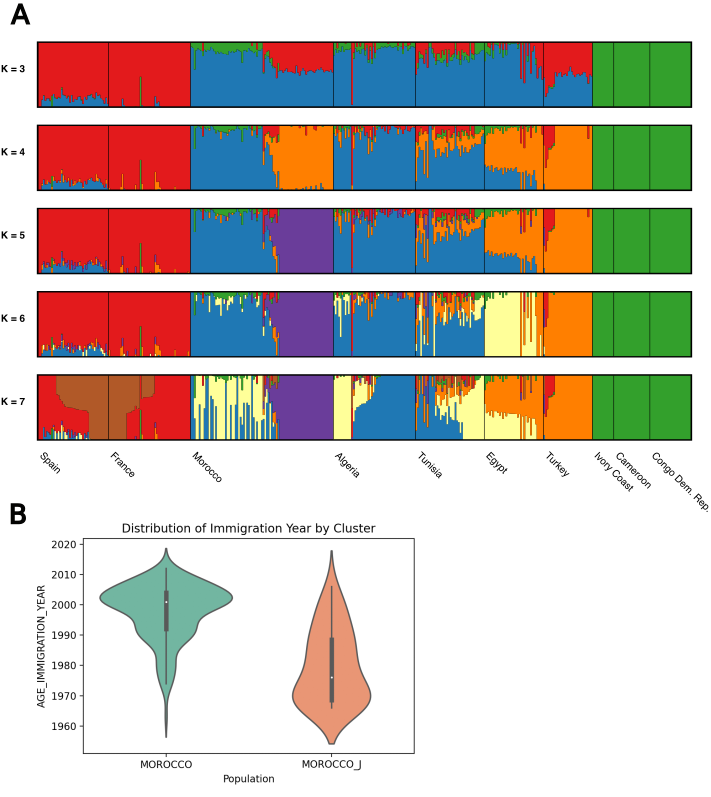

**Supplementary Figure 5. Admixture analysis of CaG participants with four grandparents born in Haiti.** (**A**) Estimation of admixture proportions among CaG participants whose four grandparents were born in Haiti (QHA) for K=3 to K=7. We included as reference CaG participants with four grandparents born within the same country in Europe, Western and Central Africa, and the Americas, and three populations from the 1000 genomes project. YRI: Yoruba in Nigeria, CHB: Han Chinese in Beijing, China, ACB: African Caribbean in Barbados. (**B**) Distribution of immigration year among QHA CaG participants born in Haiti.

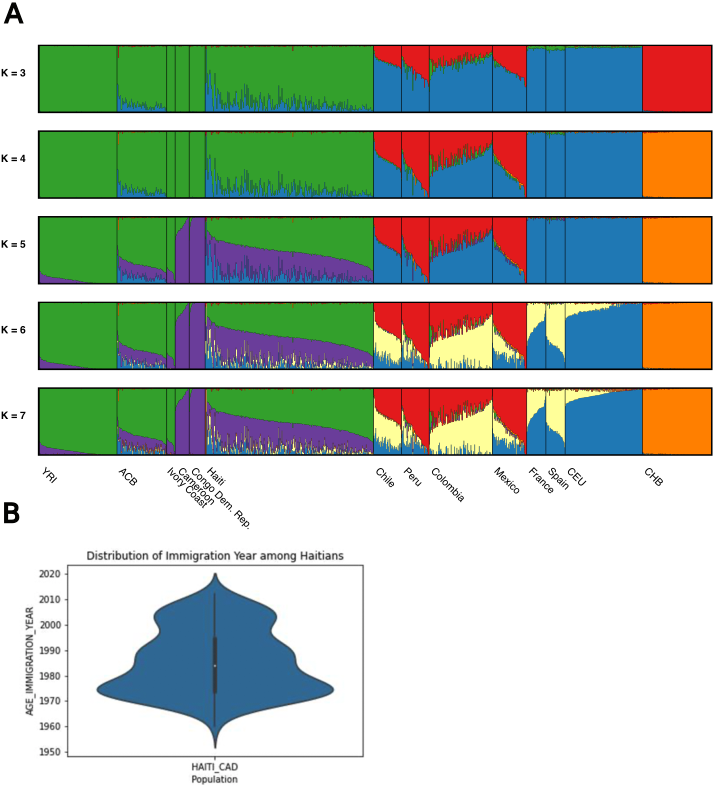

**Supplementary Figure 6.** Estimation of continental ancestry components among a random sample of 500 CaG participants whose 4 grandparents were born in Canada for K=2 to K=4. We included as reference CaG participants with four grandparents born within the same country in the Americas, Europe, and Africa.

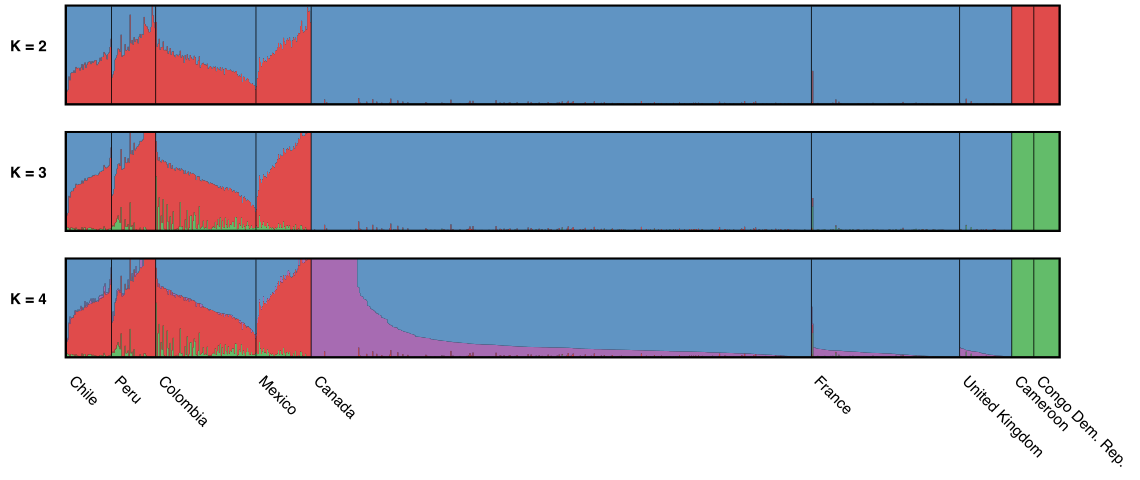

**Supplementary Figure 7.** **Population structure of the CaG participants selected for whole-genome DNA sequencing.** Uniform manifold approximation projection (UMAP) of the first five principal components calculated using the whole-genome sequence data (common variants in linkage equilibrium). The cluster at the top mostly capture individuals from the Saguenay-Lac-St-Jean region of Quebec.

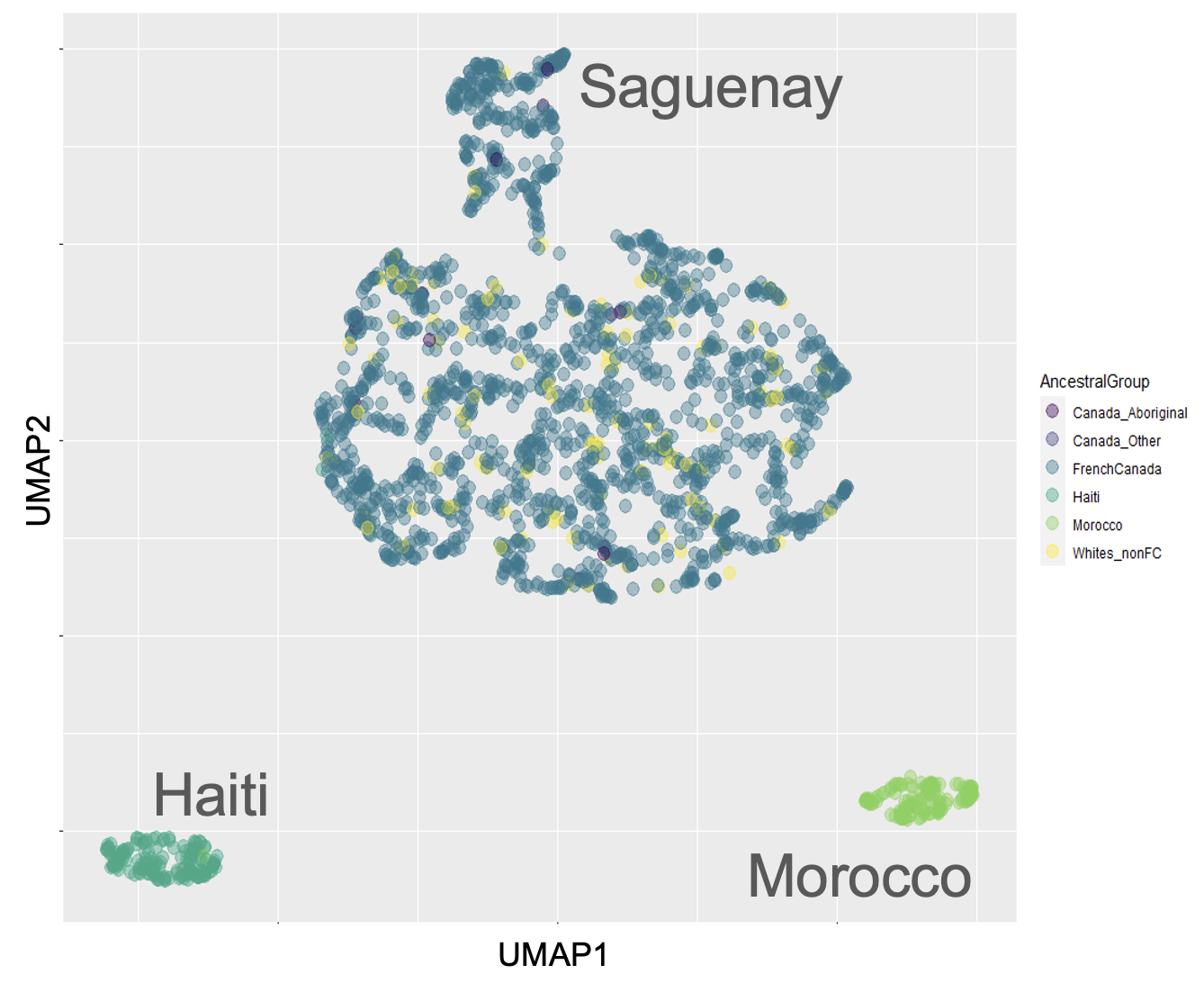

**Supplementary Figure 8. Novel variants identified by whole-genome DNA sequencing of 2,173 CARTaGENE (CaG) participants.** QFC, QMO, and QHA correspond to CaG participants with four grandparents from Canada (French Canadian), Morocco and Haiti, respectively.

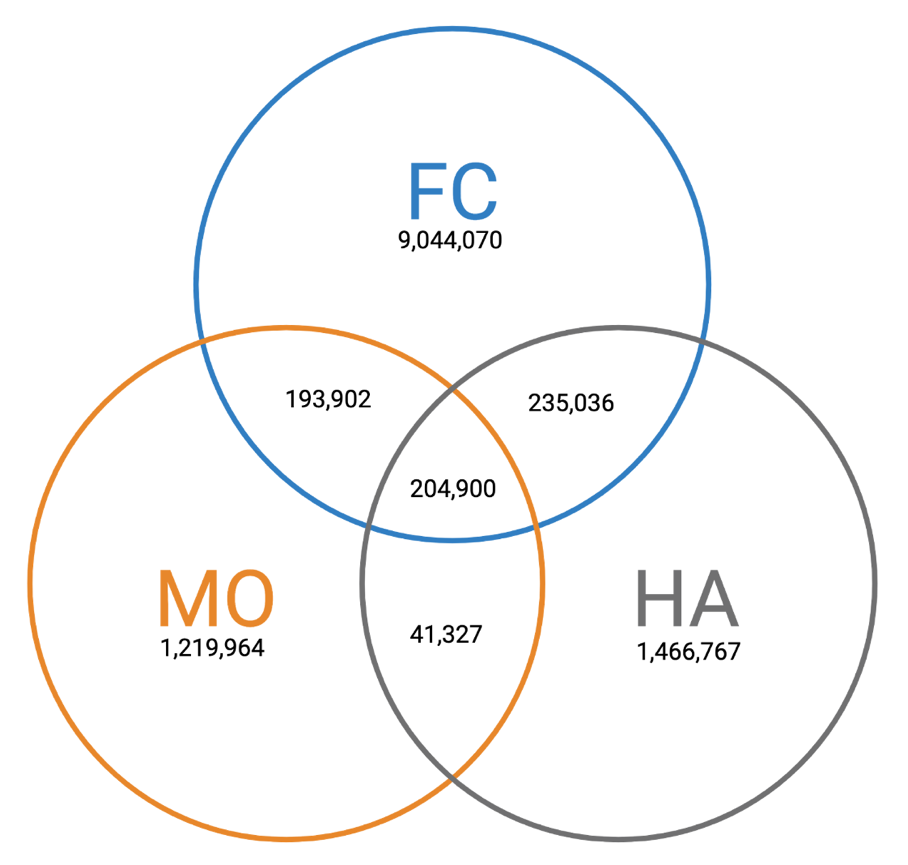

**Supplementary Figure 9. Comparison of allele frequencies for high-impact coding variants identified in CaG (y-axis) and present in the gnomAD database (x-axis).** (**A**) Allele frequencies in 1,762 French Canadians (QFC) vs. gnomAD Non-Finnish Europeans (NFE). (**B**) Allele frequencies in 163 participants of Haitian ancestry (QHA) vs. gnomAD African populations (AFR). (**C**) Allele frequencies in 132 participants of Moroccan ancestry (QMO) vs. gnomAD Middle East populations (MID). Black points represent variants annotated in ClinVar as pathogenic. Turquoise diamonds represent enriched variants (>4-times more frequent in CaG vs. gnomAD) with strong clinical evidence.

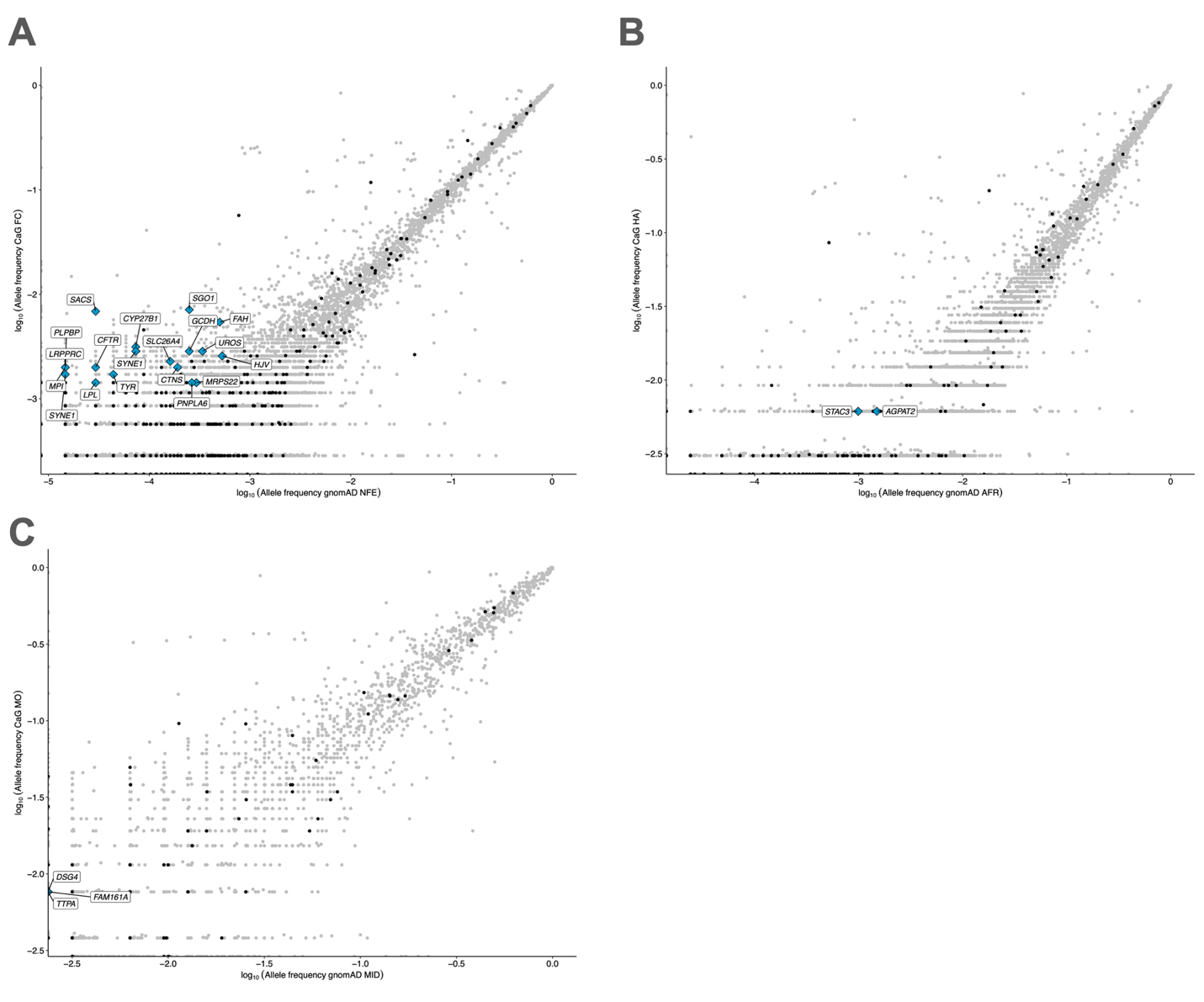

**Supplementary Figure 10. Identification of structural variants (SV) in the CARTaGENE (CaG) whole-genome DNA sequence (WGS) dataset.** (**A**) Number of SV identified per chromosome. (**B**) Distribution of the number of SV identified per genome (number of non-homozygous reference genotype per genome). (**C**) Comparison of the SV allele frequencies in CaG with the allele frequencies in gnomAD_SV. Two SVs were considered the same variant if they were of the same type and their overlap was ≥50%. (**D**) Proportion of SV of different allele frequency categories per ancestral group (All=all individuals, Can=Canadian, FC=French Canadian, H=Haitian, M=Moroccan). (**E**) Distribution of the SV lengths. (**F**) Number of SV identified in each ancestral group. (**G**) Uniform manifold approximation projection (UMAP) of the ten first principal components calculated using common SV in linkage equilibrium. (**H**) Location of the SV in the gene coding regions of the genome (UTR=SV is in the UTR of a gene with no CDS, CDS-3’UTR=the SV starts in the CDS and ends after the CDS end, CDS=SV is between the CDS start and the CDS end, 5’UTR-CDS=SV starts before the CDS start and ends in the CDS, etc.). The annotation of the SVs was generated with AnnotSV.

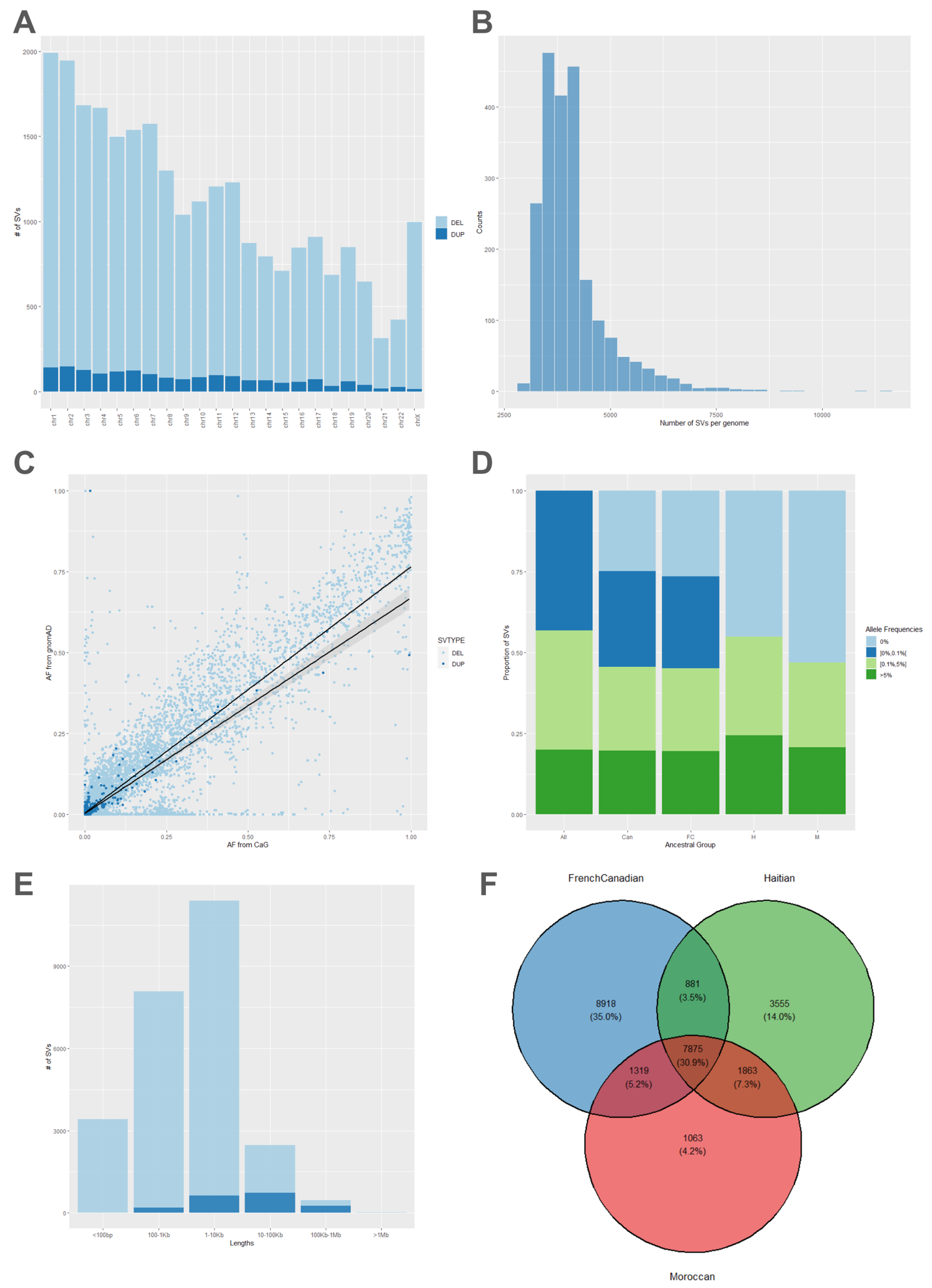

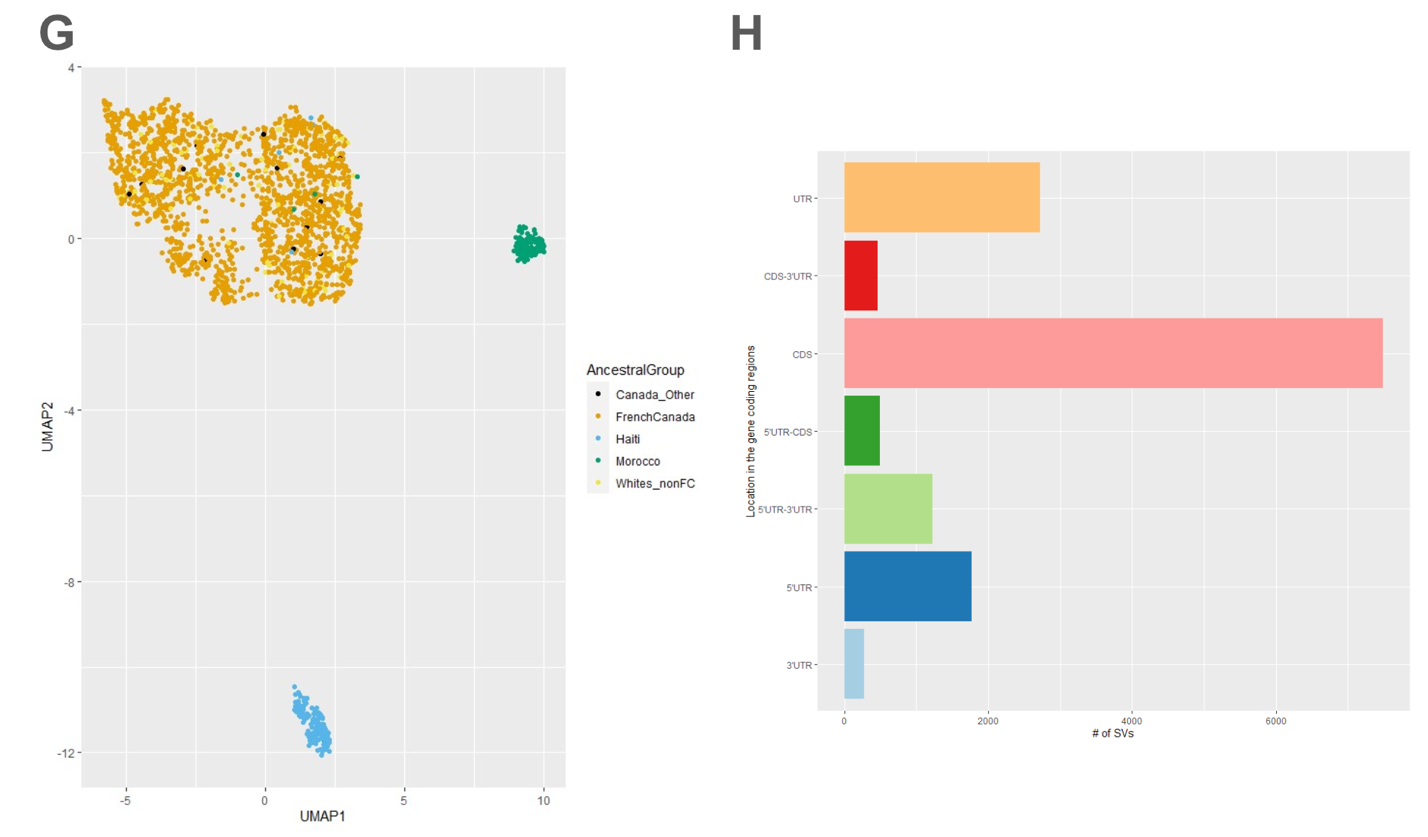

**Supplementary Figure 11. HLA allele distribution and variation in the CaG WGS cohort.** (**A**) Number of unique alleles (y-axis) observed in CaG per HLA locus (y-axis). (**B**) Low-resolution (one-field) HLA allele frequencies per region of Quebec in the CaG WGS cohort compared to regional frequencies reported in the Hema-Quebec biobank^11^. Quebec regions in CaG derived from sampling location, matched to regional allele frequency as reported in Hema-Quebec. For each region, top panel axes depict unmodified frequency, and bottom panel axes are log-scaled. Pearson R2 correlation for regional frequency between cohorts is reported for each matched region. Overall correlation between cohorts was 0.9694. (**C**) HLA allele frequencies in the CaG WGS cohort (y-axis) compared to frequencies reported in a global multi-ancestry panel (N=21,546)^8^ (x-axis). Left panel: Axes are log-scaled to resolve low-frequency alleles. Significantly enriched/different allele frequencies determined by Fisher’s Exact test (**Supplementary Table 7**). n.s.= not significant.

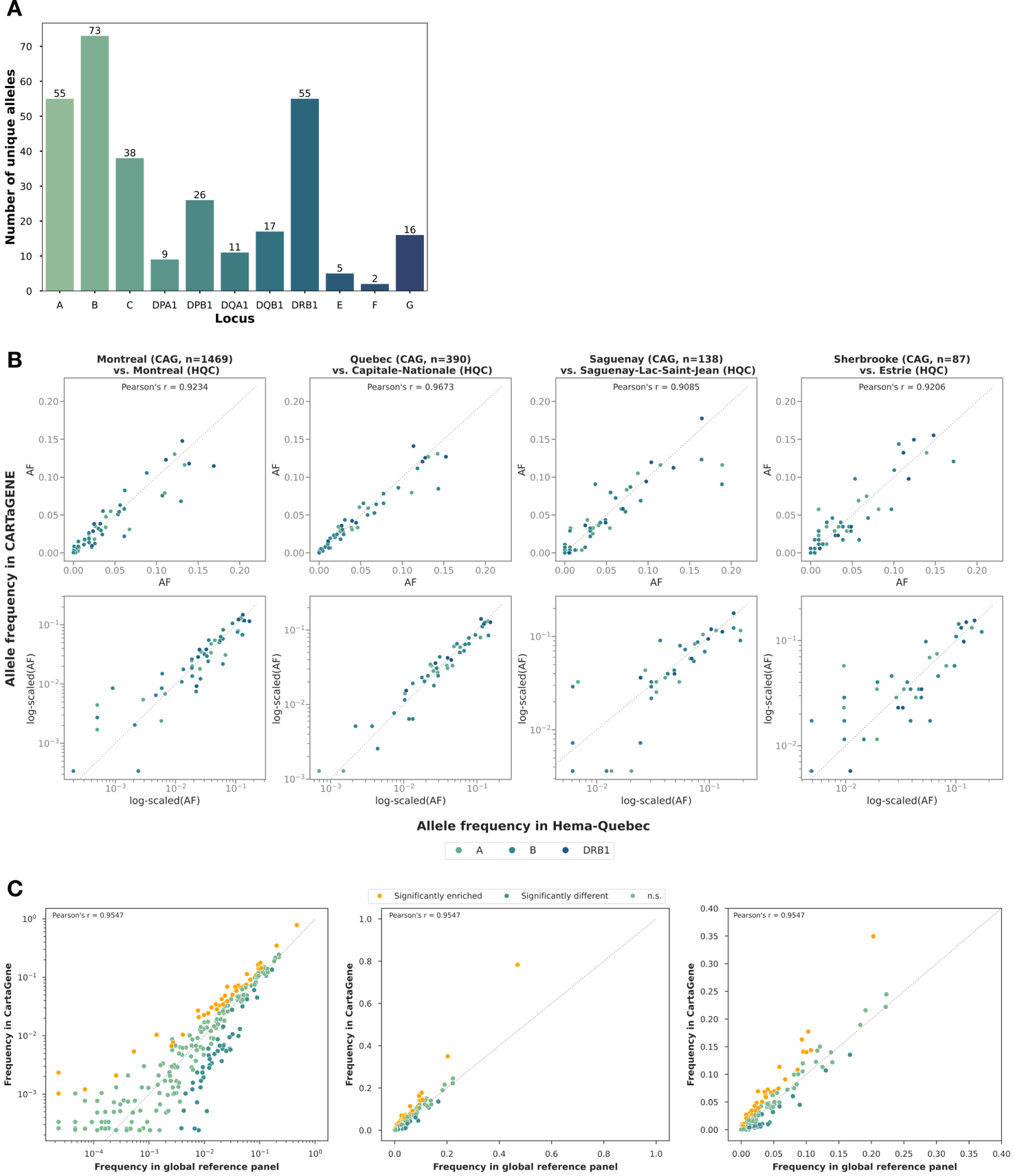

**Supplementary Figure 12. Switch error rates (SER) in 60 trios after statistical phasing together with the CaG reference panel.** AFR (N=10) - African individuals from 1000 Genomes Project. AMR (N=10) - Admixed American individuals from the 1000 Genomes Project. CSA (N=10) – Central South Asian individuals from 1000 Genomes Project. EAS (N=10) – East Asian individuals from 1000 Genomes Project. EUR (N=10) – European individuals from 1000 Genomes Project. QFC (N=10) – Quebec residents of French-Canadian ancestry. Each panel represent statistical phasing results with three different methods, from left to right: Eagle v2.4.1, Beagle v5.4, SHAPEIT5 v5.1.1. The box bounds the IQR, and Tukey-style whiskers extend to a maximum of 1.5 × IQR beyond the box. The horizontal line within the box indicates the median value. Open circles are data points corresponding to the SER in an individual. The average values and standard errors are provided in **Supplementary** **Table 10**.

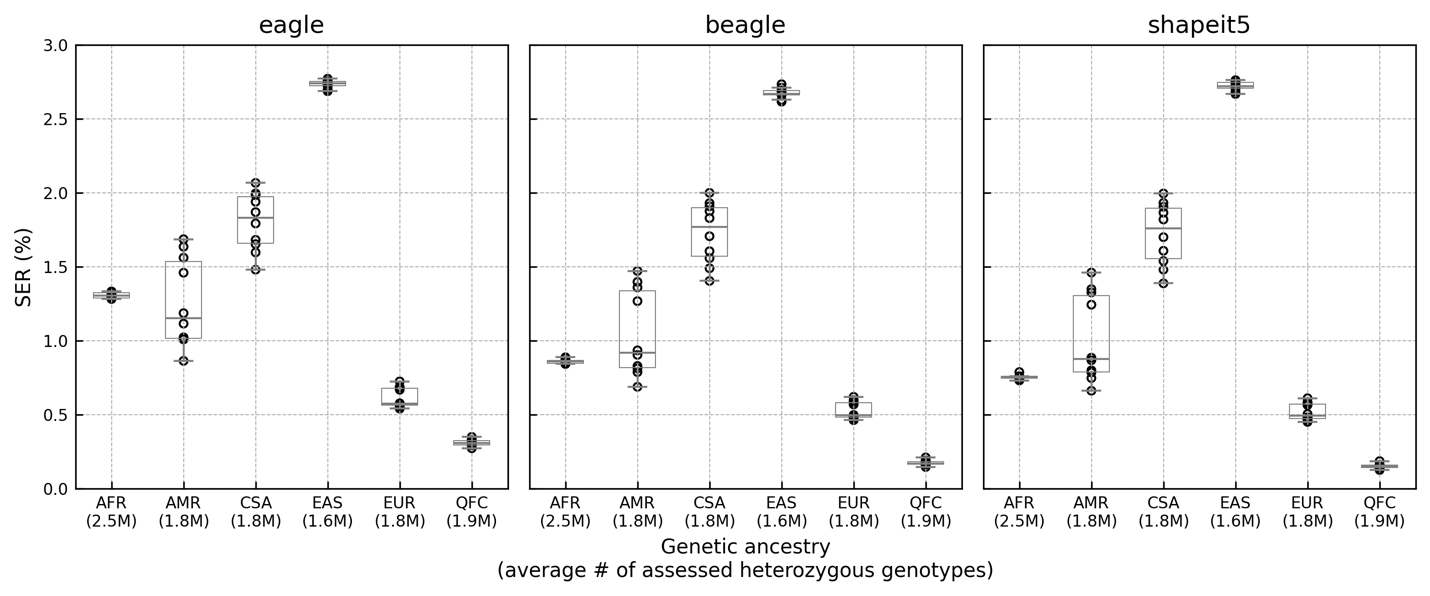

**Supplementary Figure 13. Switch error rates (SER) in 60 trios statistically phased together with the CaG reference panel without and with prior phase information from the external 1000 Genomes Project and Human Genome Diversity Project reference panels.** AFR (N=10) - African individuals from 1000 Genomes Project. AMR (N=10) - Admixed American individuals from the 1000 Genomes Project. CSA (N=10) – Central South Asian individuals from 1000 Genomes Project. EAS (N=10) – East Asian individuals from 1000 Genomes Project. EUR (N=10) – European individuals from 1000 Genomes Project. QFC (N=10) – Quebec residents of French-Canadian ancestry. The box bounds the IQR, and Tukey-style whiskers extend to a maximum of 1.5 × IQR beyond the box. The horizontal line within the box indicates the median value. (**A**) SER with and without using prior phase information from the external reference panel. The external reference panel consisted of haplotypes from sequenced individuals in 1000 Genomes Project and Human Genome Diversity Project (N=3,941 after removing trios and related individuals). The statistical phasing was performed with the SHAPEIT5 v5.1.1 as it was the only method allowing reference-based phasing while keeping variants that do not overlap between study and the reference. Open squares and diamonds are data points corresponding to the SER in an individual after statistical phasing without and with reference panel, respectively. The p-values above boxplots correspond to the one-tailed Wilcoxon signed-rank test between SER after statistical phasing without and with external reference panel. (**B**) Difference in SER between statistical phasing without and with prior information from the external reference panel. Positive values indicate that SER was lower after statistical phasing with prior information from the external reference panel. Open circles are data points corresponding to the SER difference in a single tested individual. The average values and standard errors are provided in **Supplementary Table 11**.

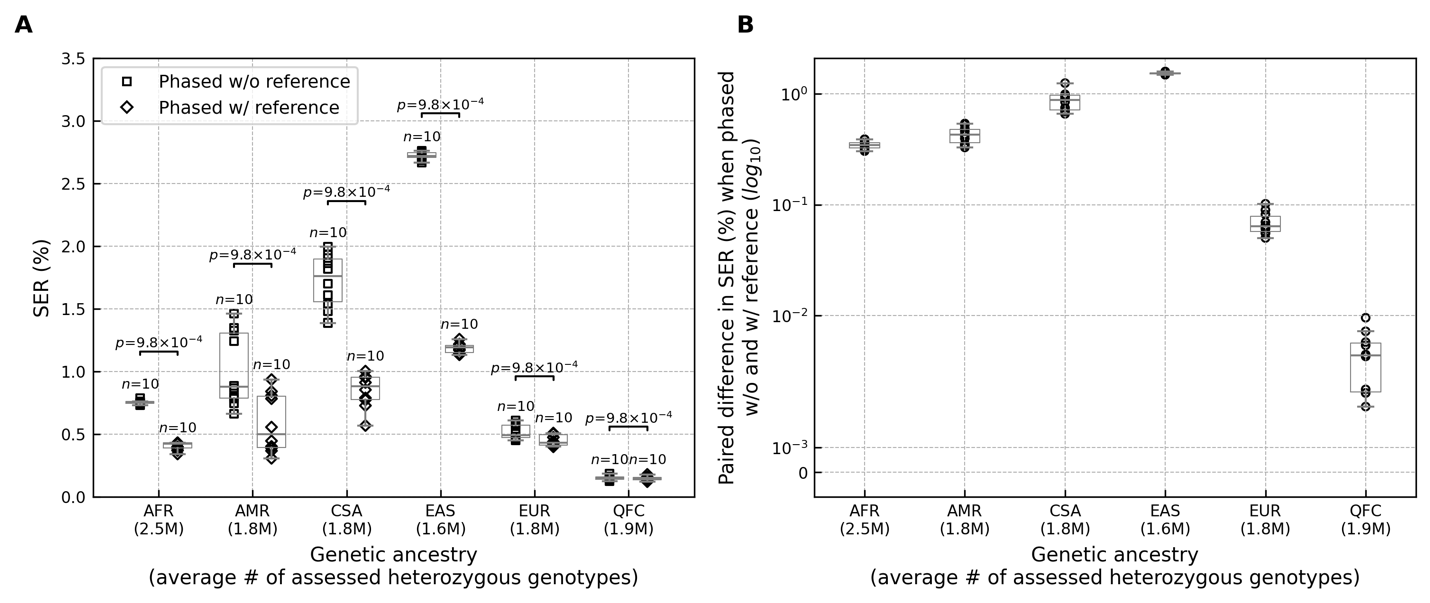

**Supplementary Figure 14. Switch error rates (SER), stratified by reference panel alternate allele count (RAC), in 60 trios statistically phased with the CaG reference panel without and with prior phase information from the external 1000 Genomes Project and Human Genome Diversity Project reference panels.** AFR (N=10) - African individuals from 1000 Genomes Project. AMR (N=10) - Admixed American individuals from the 1000 Genomes Project. CSA (N=10) – Central South Asian individuals from 1000 Genomes Project. EAS (N=10) – East Asian individuals from 1000 Genomes Project. EUR (N=10) – European individuals from 1000 Genomes Project. QFC (N=10) – Quebec residents of French-Canadian ancestry. The external reference panel consisted of haplotypes from sequenced individuals in the 1000 Genomes Project and Human Genome Diversity Project (N=3,941 after removing trios and related individuals). The statistical phasing was performed with the SHAPEIT5 v5.1.1 as it was the only method allowing reference-based phasing while keeping variants that do not overlap between the study and the reference. In **A** and **B**. Each line represents the average SER of genotypes at variants in the corresponding RAC bin. The shaded area around each line corresponds to the standard error (SE) of the average SER of genotypes at variants in the corresponding RAC bin. (**A**) Average SER of genotypes at low-frequency variants with RAC≤50. (**B**) Average SER of genotypes at common variants with RAC>50. (**C**) Each line represents the average difference in SER of genotypes at variants in the corresponding RAC bin after statistical phasing without and with the external reference panel. Positive values indicate that, on average, SER was lower after statistical phasing with prior information from the external reference panel. The shaded area around each line corresponds to the SE of the average difference in SER of genotypes at variants in the corresponding RAC bin. **Supplementary Table 12** shows the corresponding statistics and standard errors.

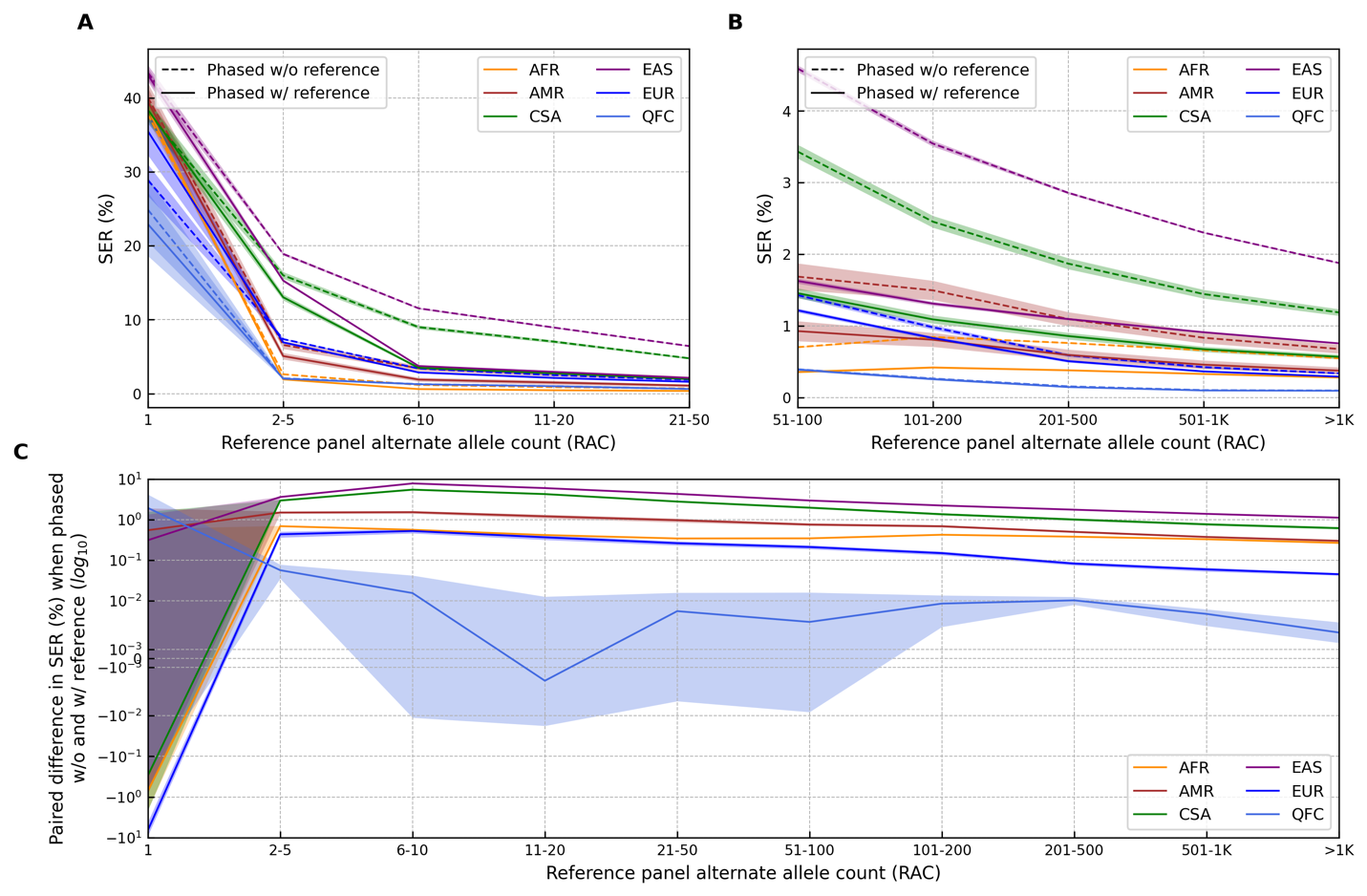

**Supplementary Figure 15. Genotype imputation using CaG and TOPMed r3 panels in 141 unrelated individuals from Quebec.** The high-depth WGS data on 141 unrelated individuals was subset to typical genotyping array positions, i.e. around 600,000 Illumina GSA v2 array positions, and imputed using two imputation panels. Only variants outside challenging regions (as defined by the Genome in a Bottle (GIAB) consortium) were used for the comparison. (**A**) The average number of variants identified per individual using high-depth WGS data, genotype imputation with CaG panel, and genotype imputation with TOPMed r3 panel. The number above each bar represents the average value, and the number in brackets below represents the standard error (SE). (**B**) Percent of imputed alternate alleles that were concordant with alternate alleles identified using high-depth WGS data. The number above each boxplot represents the corresponding average and SE (in brackets). (**C**) Fold change in the proportion of rare variants that are enriched in the QFC population in CaG. A rare variant was defined as a variant with alternate allele frequency (AF) <0.05 in the Non-Finnish European population in gnomAD v4. First, the overall proportion of rare variants that were enriched in the QFC population was computed using all variants identified in high-depth WGS data. Then, the fold change was computed with respect to the overall proportion of enriched rare variants in WGS data. Fold change equal to 1 denoted by the solid red line indicates no abundance or scarcity of enriched rare variants. AF gnomAD NFE - Allele frequency in Non-Finnish European population in gnomAD v4. QFC – Quebec residents of French-Canadian ancestry. (**D**) Median minor allele frequency (MAF) from the gnomAD v4 NFE population. The number above each boxplot represent the corresponding average of the median and SE (in brackets).

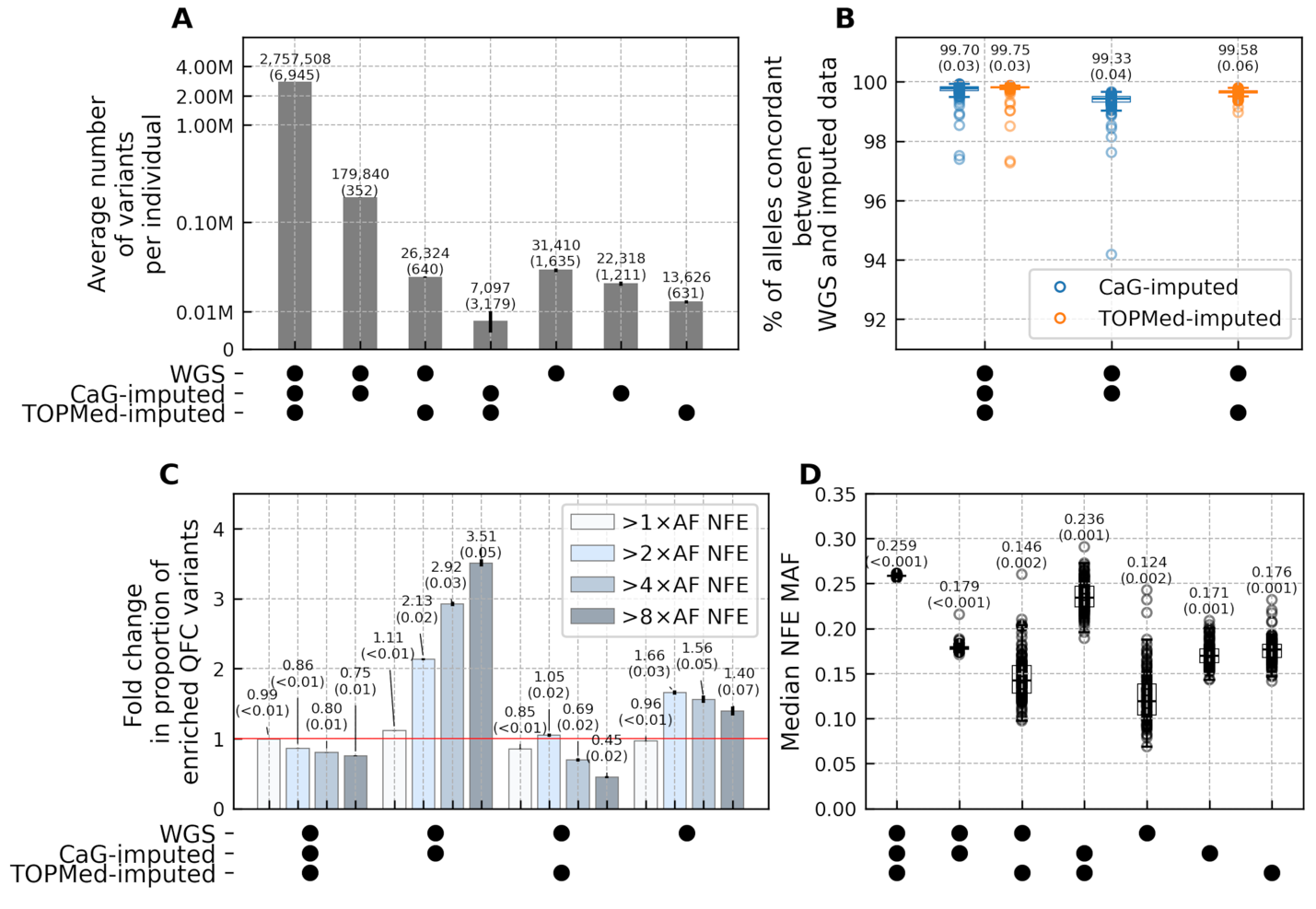

**Supplementary Figure 16. Genome-wide statistically significant loci identified using different genotype imputation approaches across all ancestry individuals in CaG.** The statistically significant independent locus was defined as the ±500-Kb region around the variant with the lowest statistically significant P-value, referred to as the lead variant. Any overlapping loci for the same trait were merged into a single locus, keeping one lead variant with the lowest P-value. Thus, by definition, each locus had only one lead variant. (**A**) The overlap between genome-wide statistically significant loci using three imputation approaches. Letters A, B, C, D, E, and F label the corresponding loci subsets. The number below the label shows the number of loci within each subset. The number in brackets corresponds to the number of lead variants not found in the alternative reference panel. (**B**) The X-axis shows the median of paired differences between imputation qualities at lead variants in CaG and TOPMed r3 imputation results. The Y axis shows the median alternate allele frequency (AF) of lead variants based on CaG imputation results. The horizontal and vertical error bars show 95% confidence intervals after 1,000,000 permutations stratified by AF. Only statistically significant (significance threshold 0.025) two-tailed permutation P-values for the median of paired differences in imputation qualities are displayed below the subset labels to reduce image cluttering. (**C**) Each bar shows the number of unique lead variants (Y-axis) within each subset in GWASs of ALL participants and participants of European-genetic ancestry only (EUR). The blue-coloured bars correspond to the number of lead variants private to QHA and QMO individuals in CaG WGS data. (**D**) The X-axis shows the median of paired differences between imputation qualities at lead variants in CaG and TOPMed r3 imputation results. The blue-coloured circles correspond to the lead variants private to QHA and QMO individuals in CaG WGS data, while grey-coloured circles correspond to all other lead variants. The horizontal box bounds the IQR, and Tukey-style whiskers extend (the black horizontal lines) to a maximum of 1.5 × IQR beyond the box.

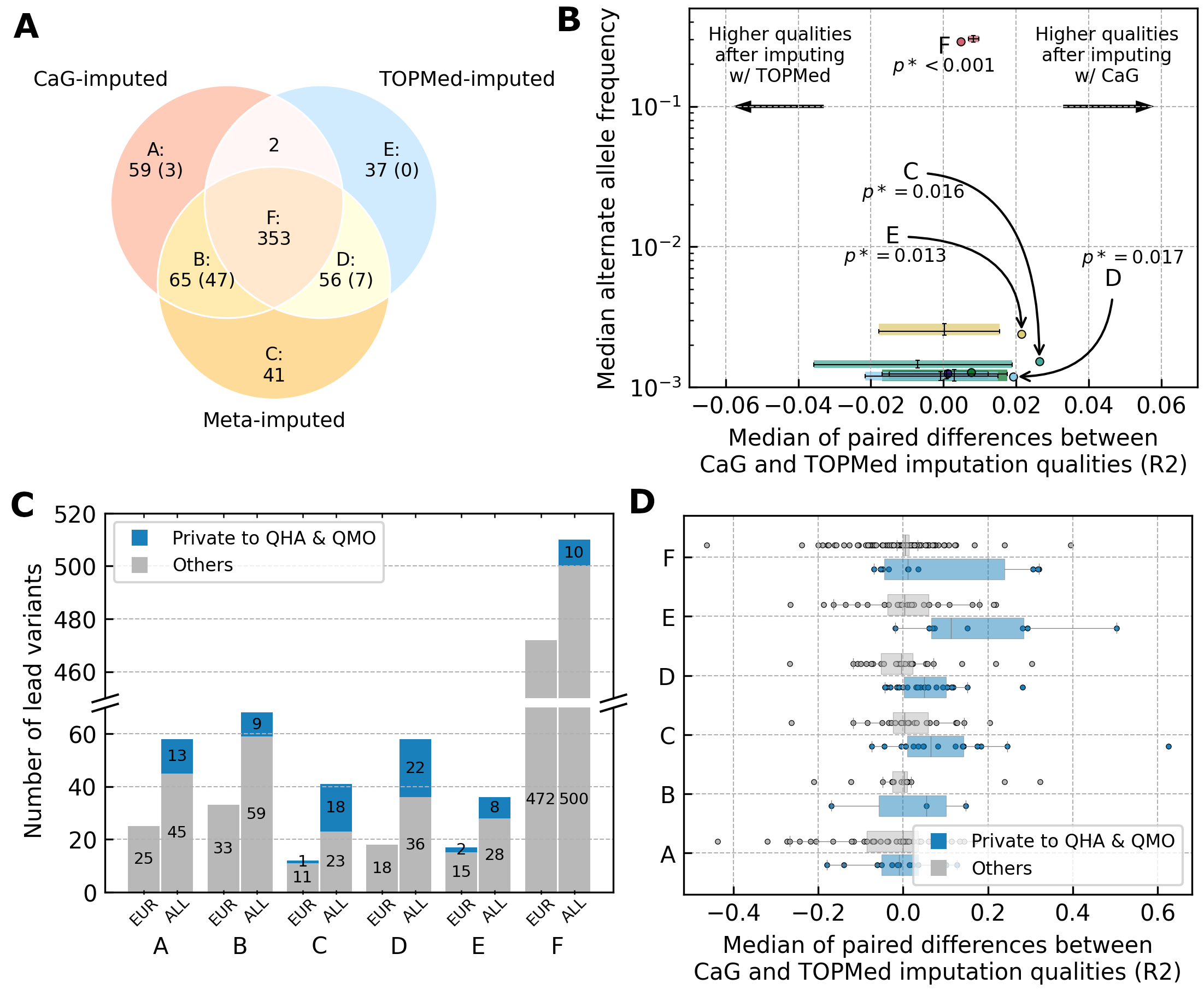

**Supplementary Figure 17. Genotype imputation qualities of variants imputed from CaG and TOPMed r3 reference panel into the CaG genotyping array data.** The figure shows genetic variants shared in CaG and TOPMed r3 reference panels. We grouped shared genetic variants into five groups (X-axis) using their alternate allele frequencies (AF) of the imputed genotypes from the CaG reference panel (we observed similar results when using AF of the genotypes imputed from the TOPMed r3 reference panel). The annotated number above each violin plot indicates the median (M) value. The black vertical box bounds the IQR, and Tukey-style whiskers extend (the black vertical lines) to a maximum of 1.5 × IQR beyond the box. The white circle within the box indicates the median value. **(A)** The distribution of the R2 imputation qualities in CaG (blue) and TOPMed r3 (orange) imputation results. **(B)** The distribution of paired differences in R2 between CaG and TOPMed r3 imputation results.

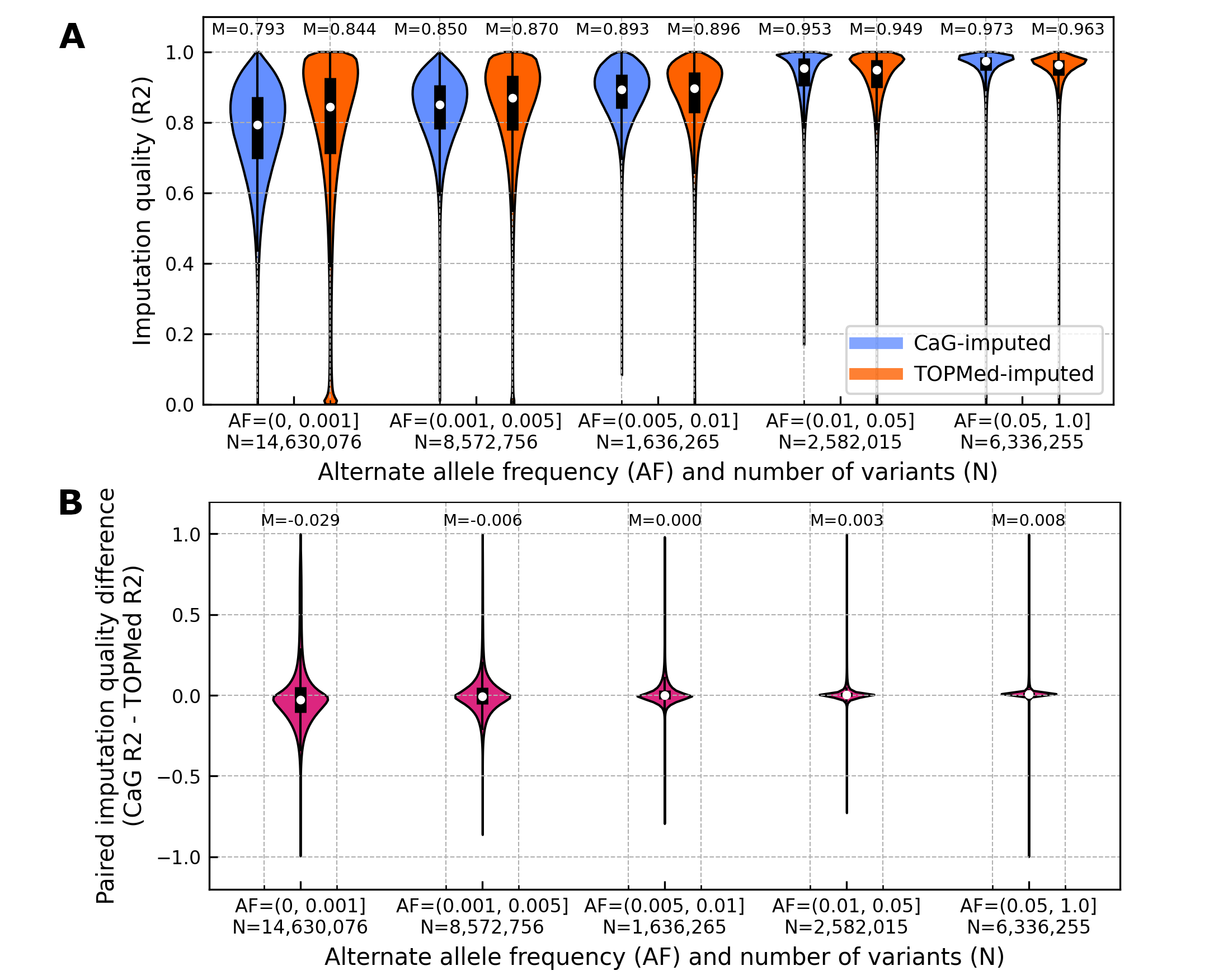

**Supplementary Figure 18. HLA allele frequencies and HLA phenome-wide association results. Top association hit, HLA-B*08:01, is enriched in the CaG WGS cohort.** (**A**) HLA-wide Manhattan plot of association results for the top phenotype, white blood cell (WBC) count, after analysis of 42 continuous traits with imputed HLA alleles using REGENIE. X-axis indicates position along the HLA region, points are coloured by HLA locus. Y-axis depicts negative log-scaled P-value. Significant hits after correction for multiple testing (threshold indicated at grey dotted line; P-value <3.7x10^-4^) are labelled. Association results are from the entire imputed CaG cohort (multi-ancestry). (**B**). PheWAS “Miami” plot depicting association results across entire spectrum of phenotype traits analysed for top hit, HLA-B*08:01. Top panel: Association results in entire imputed CaG cohort (multi-ancestry). Y-axis depicts negative log-scaled P-values. Bottom panel: Association results in EUR-ancestry subset of CaG imputed cohort. Y-axis depicts negative log-scaled P-values (axis flipped). X-axis indicates abbreviated phenotype label; phenotype names are given in full in legend below. Marker arrow direction indicates direction of effect of association.

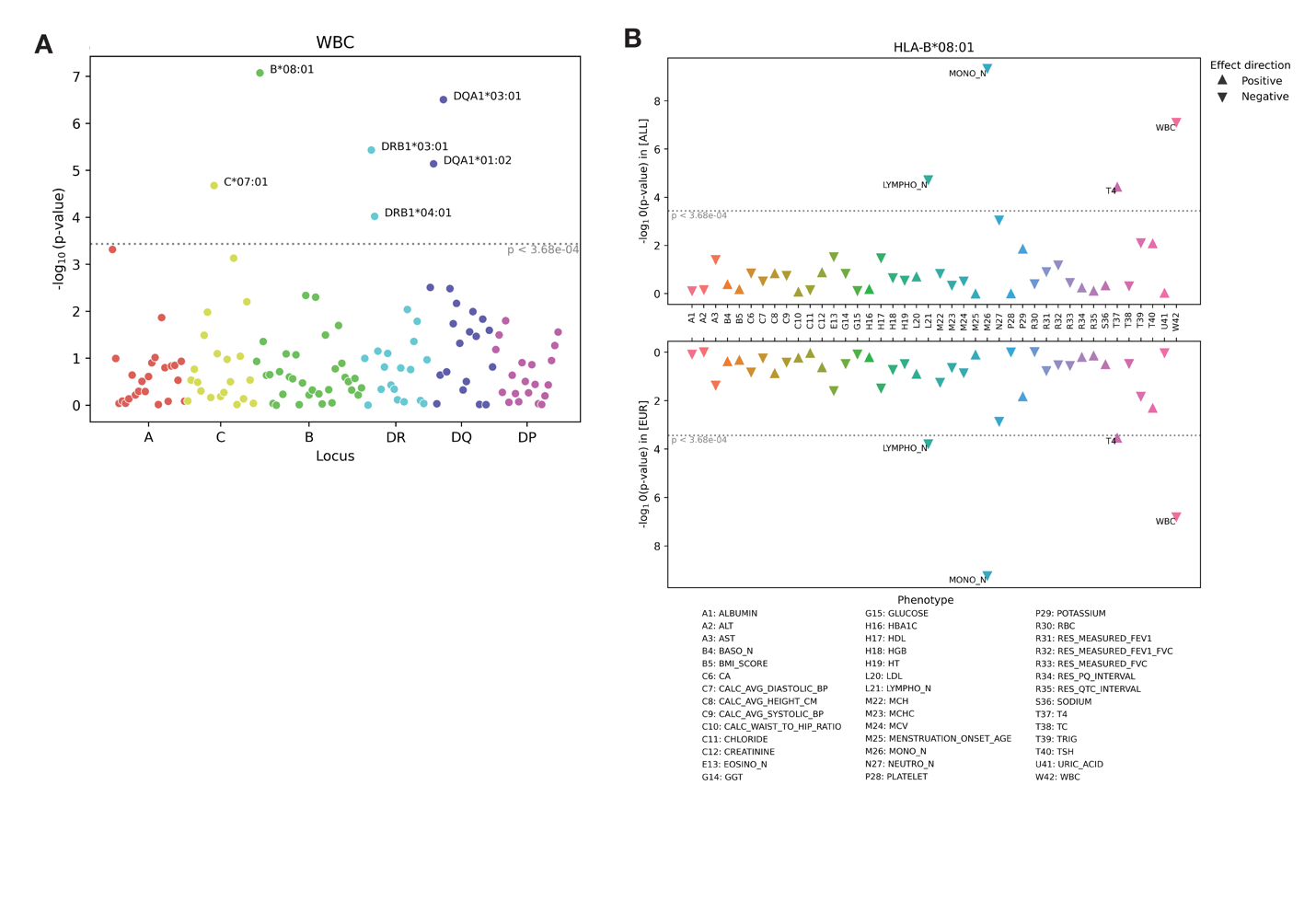

**Supplementary Figure 19. Association results in *INSR* with TSH levels.** The linkage disequilibrium (LD) r^2^ metric between the lead variant and other variants was computed based on the imputed dosages using the corresponding reference panel. The LD metrics are visualized relative to the variant represented by the diamond shape. The up-pointing triangles represent positive effect sizes, while the down-pointing triangles represent the negative effect size. (**A**) Genetic association results after imputation with the CaG panel. The lead variant is rs796144540 (19:7238617:CA/C) and is represented by the diamond shape. (**B**) Genetic association results after imputation with the CaG panel and after conditioning on the rs796144540 variant. The lead variant is rs8107575 (19:7165528:C/T) is represented by the diamond shape. (**C**) Genetic association results after imputation with the TOPMed r3 panel. The diamond shape represents rs8107575 (19:7220585:G/T).

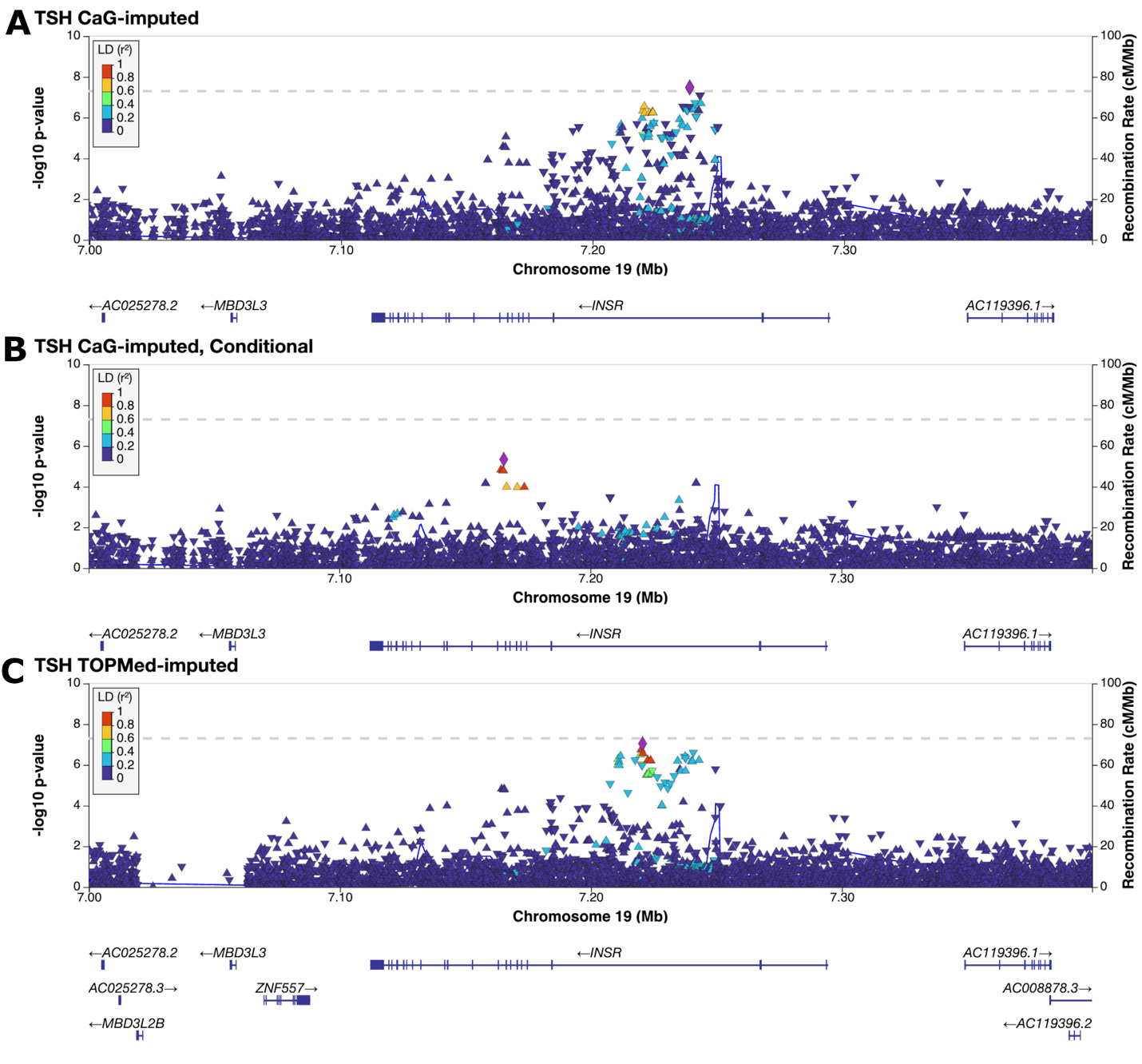

**Supplementary Figure 20. Association results in *PDE6D* locus with basophils count.** The linkage disequilibrium (LD) r^2^ metric between the lead variant and other variants was computed based on the imputed dosages using the corresponding reference panel. The LD metrics are visualized relative to the variant represented by the diamond shape. The up-pointing triangles represent positive effect sizes, while the down-pointing triangles represent the negative effect size. (**A**) Genetic association results after imputation with the CaG panel. The lead variant is rs867418280 (2:231832599:C/T) and is represented by the diamond shape. (**B**) Genetic association results after imputation with the CaG panel and after conditioning on the rs867418280 variant. The lead variant is rs1187982745 (2:231767325:G/GT) and is represented by the diamond shape. (**C**) Genetic association results after imputation with the TOPMed r3 panel. The diamond shape represents rs9967791 (2:231821814:C/G).

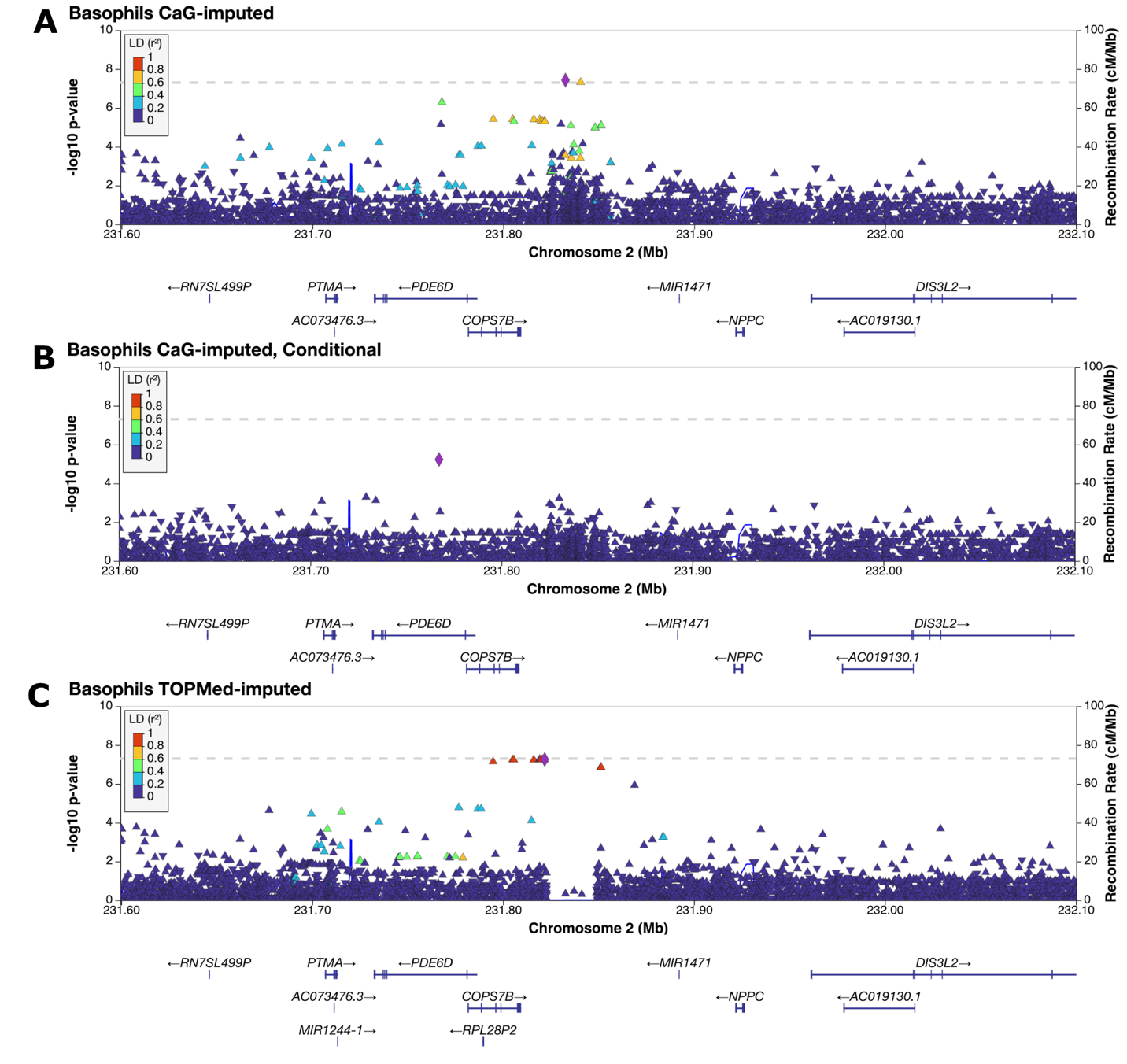

**Supplementary Figure 21. Imputation quality for prioritized candidate functional coding and non-coding variants.** Imputation quality for enriched pathogenic/high-impact, conservation or epigenetics variants (see **Methods** for variant definition based on FAVOR annotations) in CARTaGENE European-ancestry participants using different imputation panels. The percentages are the proportion of variants with poor imputation quality (rsq_hat <0.3). (**A**) Violin plots for all variants. (**B**) Zoom-in of the distributions for the pathogenic/high-impact variants. (**C**) Upset plots that shows the number of candidate functional variants that are well-imputed (rsq_hat <0.3) using any of the three imputation panels. Using the CaG-only imputation panel or the meta-imputation approach yield the same variants (99.95%), but the imputation quality is slightly higher with meta-imputation (see **A** and **B**).

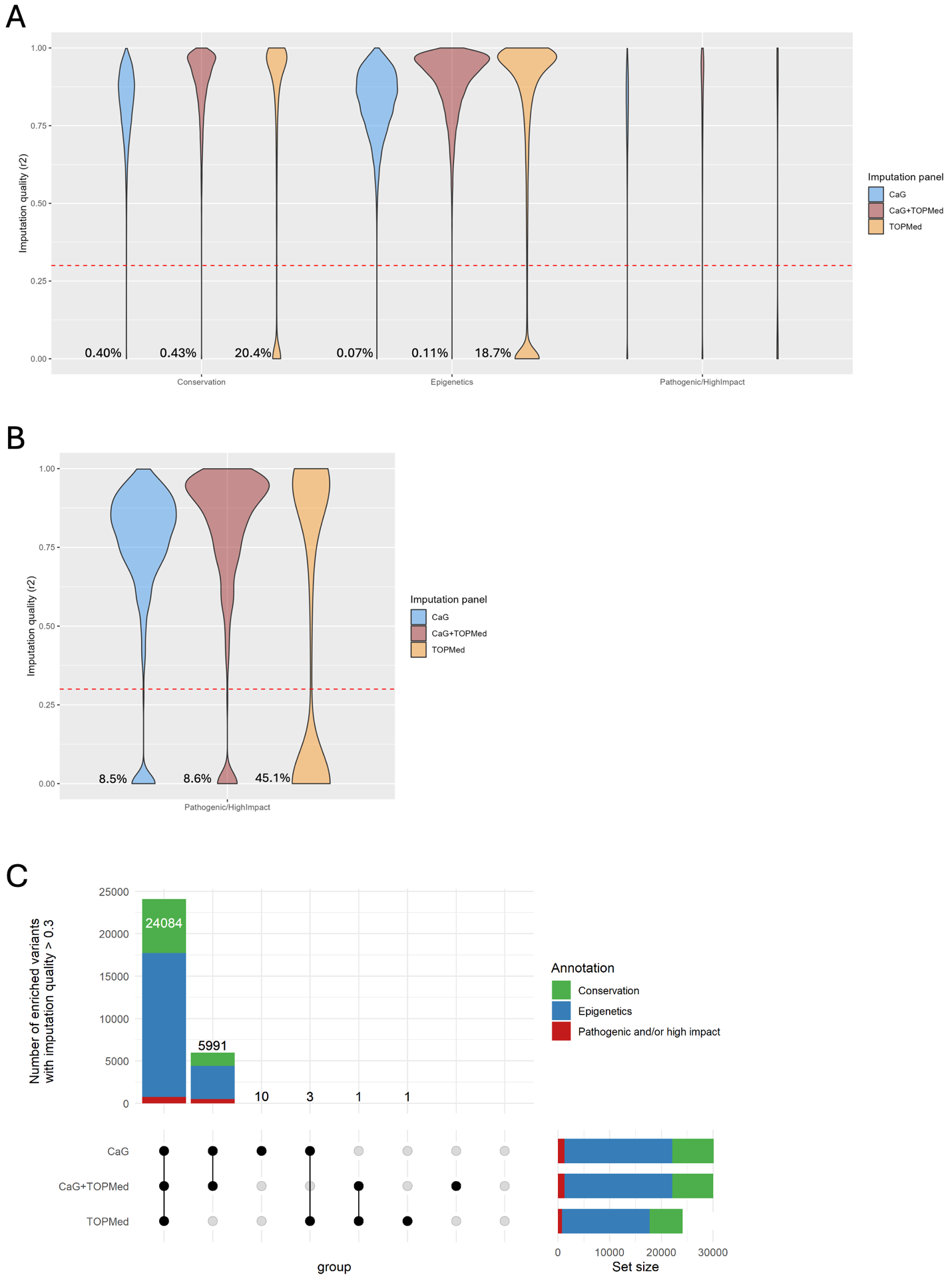

**Supplementary Figure 22. eQTL analyses in CaG.** (**A**) Correlation between effect sizes for variant-gene eQTL associations tested in CaG and significant associations in GTEx whole-blood eQTL (Spearman’s correlation of 0.82 and P-value <2.2x10^-16^). (**B**) Comparison of the median TPM values, in log scale, between GTEx Whole blood and CaG RNA-sequencing experiments.

**A**

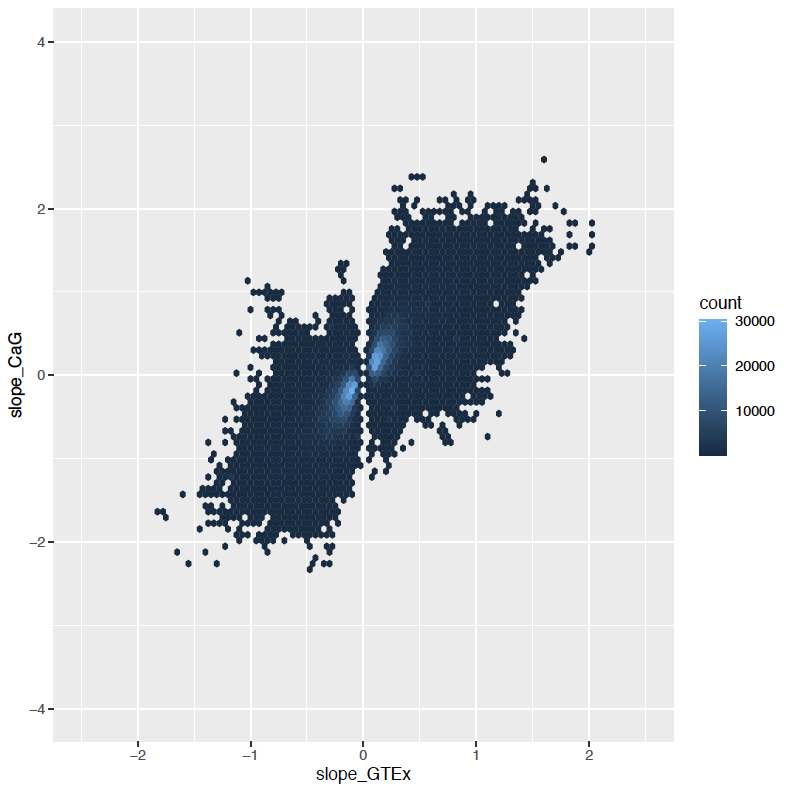

**B**

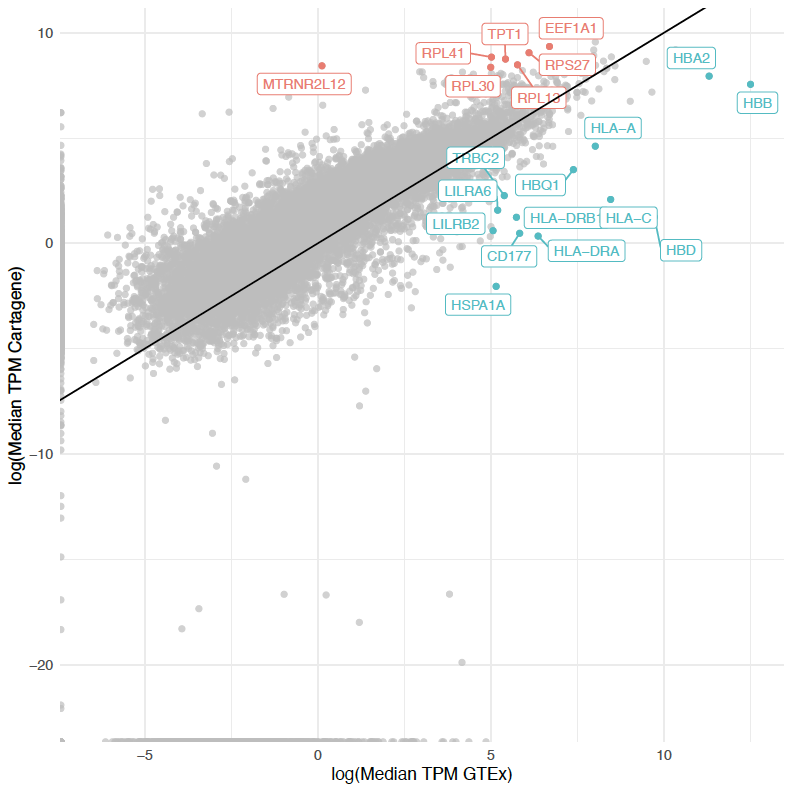
